# Objective evaluation of clinical actionability for genes involved in myopathies: 63 genes with a medical value for patient care

**DOI:** 10.1101/2022.05.16.22275131

**Authors:** M. Vecten, E. Pion, M. Bartoli, R. Juntas Morales, D. Sternberg, J. Rendu, T. Stojkovic, C. Acquaviva Bourdain, C. Métay, I. Richard, M. Cerino, M. Milh, S. Gorokhova, N. Levy, X. Latypova, G. Bonne, V. Biancalana, F. Petit, A. Molon, A. Perrin, P. Laforet, S. Attarian, M. Cossée, M. Krahn

## Abstract

The implementation of high-throughput diagnostic sequencing has led to the generation of large amounts of mutational data, making their interpretation more complex and responsible for long delays. It has been important to prioritize certain analyses, particularly those of “actionable” genes in diagnostic situations, involving specific treatment and/or management. In our project, we carried out an objective assessment of the clinical actionability of genes involved in myopathies, for which only few data obtained methodologically exist to date. Using the ClinGen Actionability criteria, we scored the clinical actionability of all 199 genes implicated in myopathies published by FILNEMUS for the “National French consensus on gene Lists for the diagnosis of myopathies using next generation sequencing”. We objectified that 63 myopathy genes were actionable with currently available data. Among the 36 myopathy genes with highest actionability scores, only 8 had been scored to date by ClinGen. The data obtained through these methodological tools are an important resource for strategic choices in diagnostic approaches and the management of genetic myopathies. The clinical actionability of genes has to be considered as an evolving concept, in relation to progresses in disease knowledge and therapeutic approaches.

---

High-throughput sequencing revolutionized the possibilities of genetic analysis by allowing the simultaneous mutational screening of several genes, progressively extending to all the genes through exome and genome sequencing^1^. These technologies lead to the potential for the recognition of secondary findings unrelated to the indication for ordering the sequencing, but of medical value for patient care^2^. The concept of “actionable genes” has emerged in this context for diagnostic purposes. In 2013, the ACMG drafted initial recommendations on the principle of actionability and published a list of 59 actionable genes^2^. More recently, the ClinGen Actionability Working Group (AWG) proposed a semiquantitative metric scoring to assess the clinical actionability of genes through four indicators: severity of disease, penetrance/likelihood of disease, effectiveness of intervention and nature of intervention (https://www.clinicalgenome.org/site/assets/files/2180/actionability_sq_metric.png)^3^, combined with indication of the level of evidence, to aid to determine best practices regarding secondary findings^4^. Across these topics the ClinGen AWG scored 213 outcome-intervention pairs from 127 genes associated with 78 disorders^4^. This semi-quantitative measure is set to evolve thanks to the contribution of the entire international community and should be considered as a starting point in the standardization of clinical actionability.

In France, FILNEMUS was the first to publish the “gene-disease” correlations for 199 genes implicated in myopathies^5^ using the procedure published by the ClinGen Clinical Validity framework, in order to simplify the molecular diagnosis of myopathies^6^. Despite this, the use of next-generation-sequencing is faced with results delays, which is problematic for genes with medical value for patient care. ClinGen AWG listed some myopathy genes as actionable, such as the *GAA* gene responsible for Pompe disease, for which the prognosis may improve if the patient is treated quickly especially in its infantile form^7^. However, actionability has not been assessed for all myopathy genes. Based on the ClinGen AWG recommendations, we carried out a clinical actionability objective assessment of the 199 genes implicated in myopathies reported on the FILNEMUS list.

## METHODS

We used the semi-quantitative actionability metric established by ClinGen^3,4^ to score the clinical actionability of 199 genes implicated in myopathies published by FILNEMUS.^5^ The 199 genes are associated with 223 disorders. We choose to offer an overall score for each gene-associated disorder, to mark the most severe phenotype and not each symptom (cardiac involvement, muscle damage, etc.). The four indicators, Severity of disease, Penetrance/Likelihood of disease, Effectiveness of specific intervention, Nature of intervention, were each scored from 0 to 3, from a low level to a high level of actionability.^3^ For example the score Severity is 0 if the disease has minimal health impact or no morbidity and 3 if there is sudden death or inevitable death, the score Penetrance is 0 if <1% chance or unknown, 3 if >40% chance, Effectiveness of specific intervention is 0 if ineffective/no intervention, 3 if highly effective, Nature of intervention is 0 if high risk/poorly acceptable/intensive or no intervention, as 3 if low risk/medically acceptable/low intensity intervention ^3,4^. All scores have been reviewed by the French FILNEMUS expert-network (clinicians and geneticists).

We then compared the scores obtained for the myopathy genes/disorders with those of the ClinGen AWG (127 genes corresponding to 78 disorders with 213 outcome-intervention pairs)^4^, using the nonparametric Kruskal-Wallis test. Thereafter, we analysed the distribution of scores of the four indicators for the ClinGen AWG and myopathy genes using the chi-square method that admits differences in group size.

## RESULTS

The 223 myopathy genes/disorders were scored from 1 to 12 (Supplemental material 1). More than 45% of the scores are distributed between 4 and 6, while the scores for genes/disorders of the ClinGen AWG genes list were between 5 and 12, with > 50% of them greater than or equal to 9 (fig 1a).

**Figure 1.**
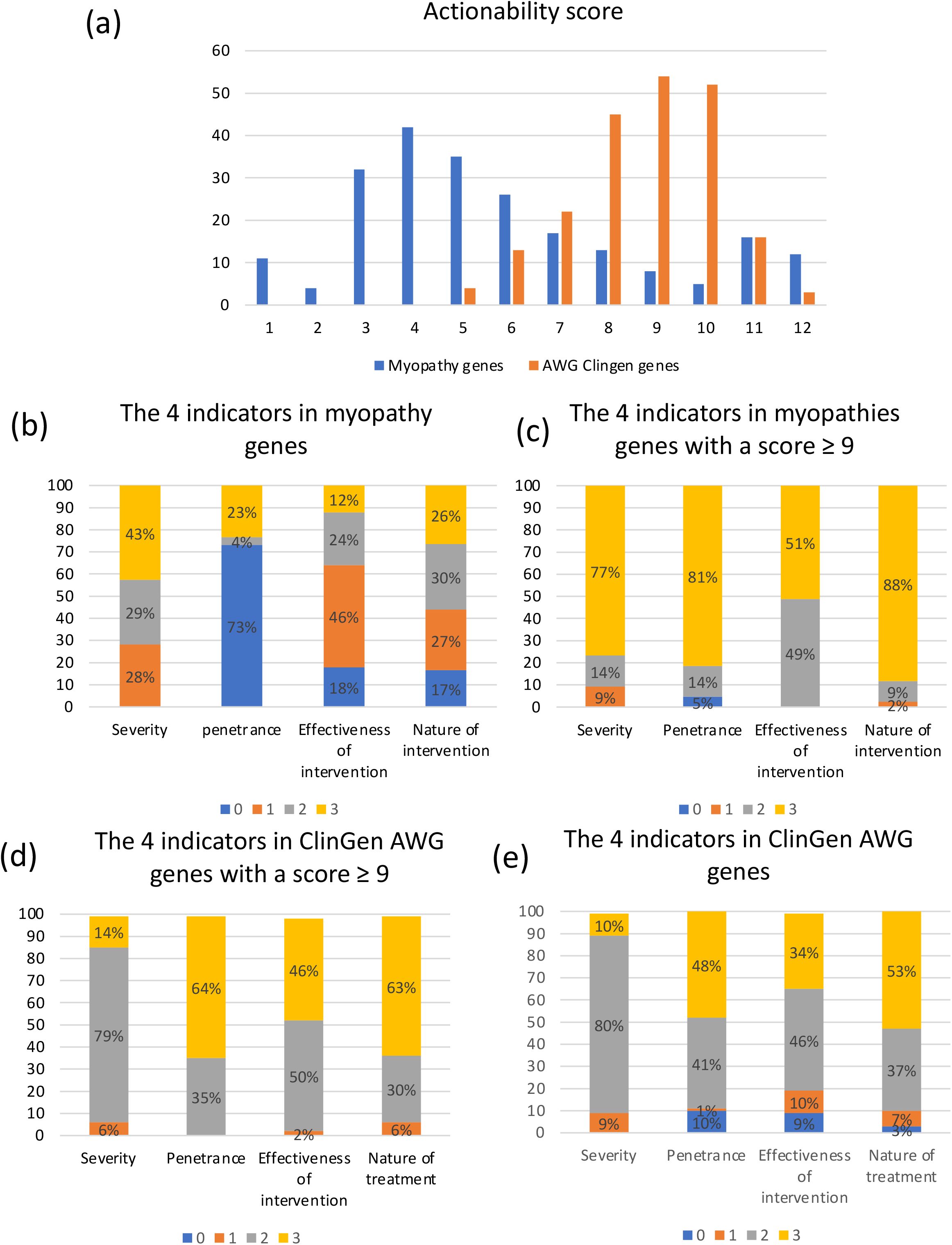
Actionability scores for myopathy genes and ACMG genes. The four indicators of actionability are Severity of disease, Penetrance/Likelihood of disease, Effectiveness of specific intervention, Nature of intervention. These indicators are each scored from 0 to 3, from a low level to a high level of actionability^3^. a/ Total scores for the 223 myopathy genes/disorders (from the 199 genes reported by FILNEMUS^5^) and for the 213 ClinGen AWG genes/outcome-intervention pairs (from the 127 genes scored by ClinGen AWG^4^). b/ Distribution of the four indicators for the 223 myopathy genes/disorders c/ Distribution of the four indicators for the 213 ClinGen AWG genes/outcome-intervention pairs d/ Distribution of the four indicators for the 43 myopathy genes/disorders (36 genes) with a global score >9. e/ Distribution of the four indicators for the 125 ClinGen AWG genes/outcome-intervention pairs (111 genes) with a global score >9.

We evaluated the scores distribution of the 4 indicators for the myopathy genes/disorders (fig 1b). Concerning the severity indicator: 43% had very high mortality rates (score 3), 29% had moderate severity (score 2) and 28% low severity (score 1). For the disease penetrance indicator (likelihood): the information was available for only 27% of the myopathy genes/disorders, leading to an artificial score of 0 for 73% of them. For the effectiveness of intervention: 36% were highly or moderate effective (score ≥2). The nature of intervention is evenly distributed between the different scores, with a great or high risk (scores 0 and 1) in 44% of cases.

By filtering on the two care indicators, effectiveness of specific intervention with a score ≥2 and nature of intervention ≥1, we retained 63 myopathy genes corresponding to 78 disorders that could be actionable on their management (table 1). Final scores of actionability of these genes/disorders were between 5 and 12, with the majority ≥9 (43/78, 55%). The intervention included either the availability of drugs or a specific cardiac management (defibrillator/pacemaker).

**Table 1:**
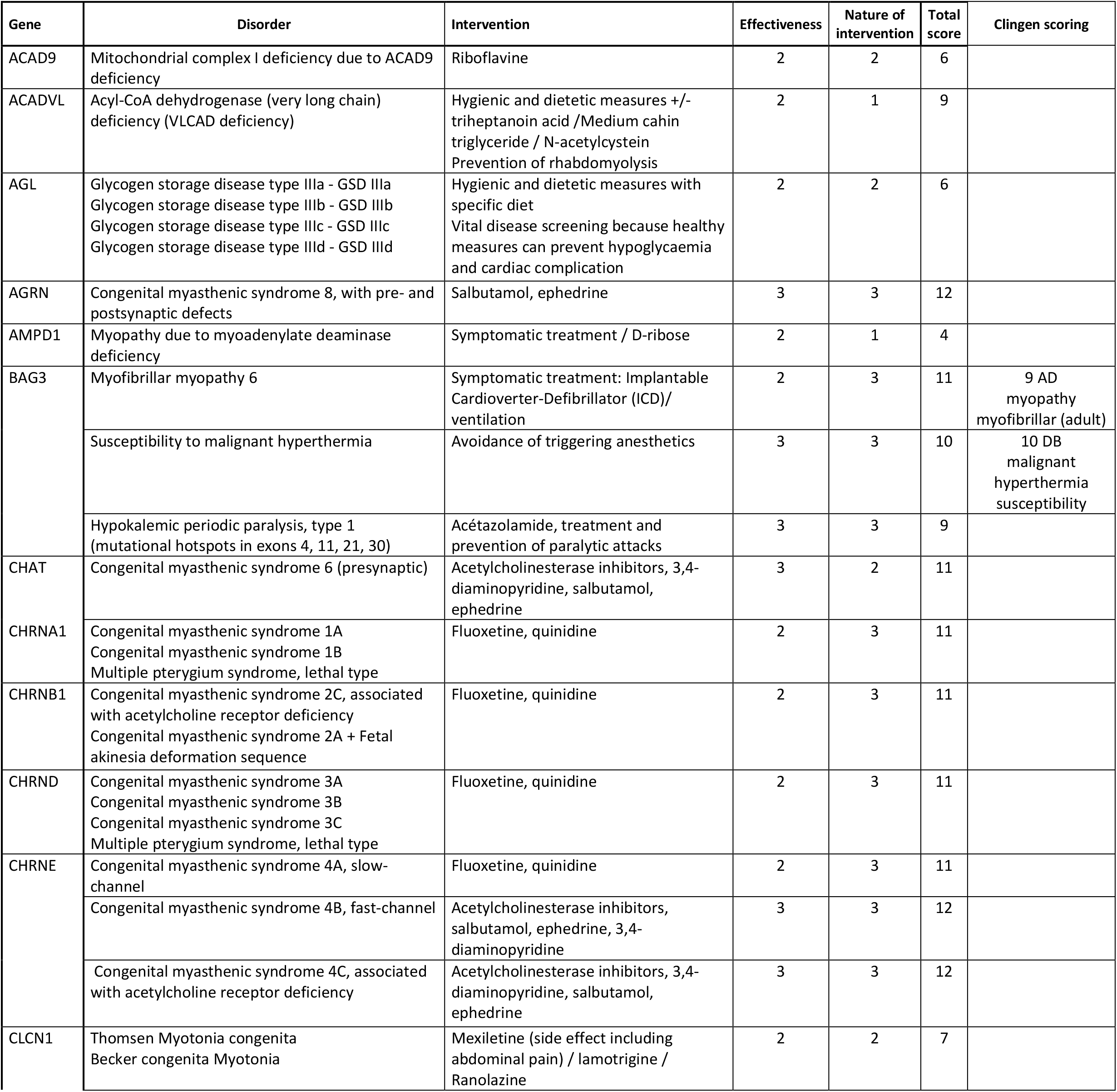
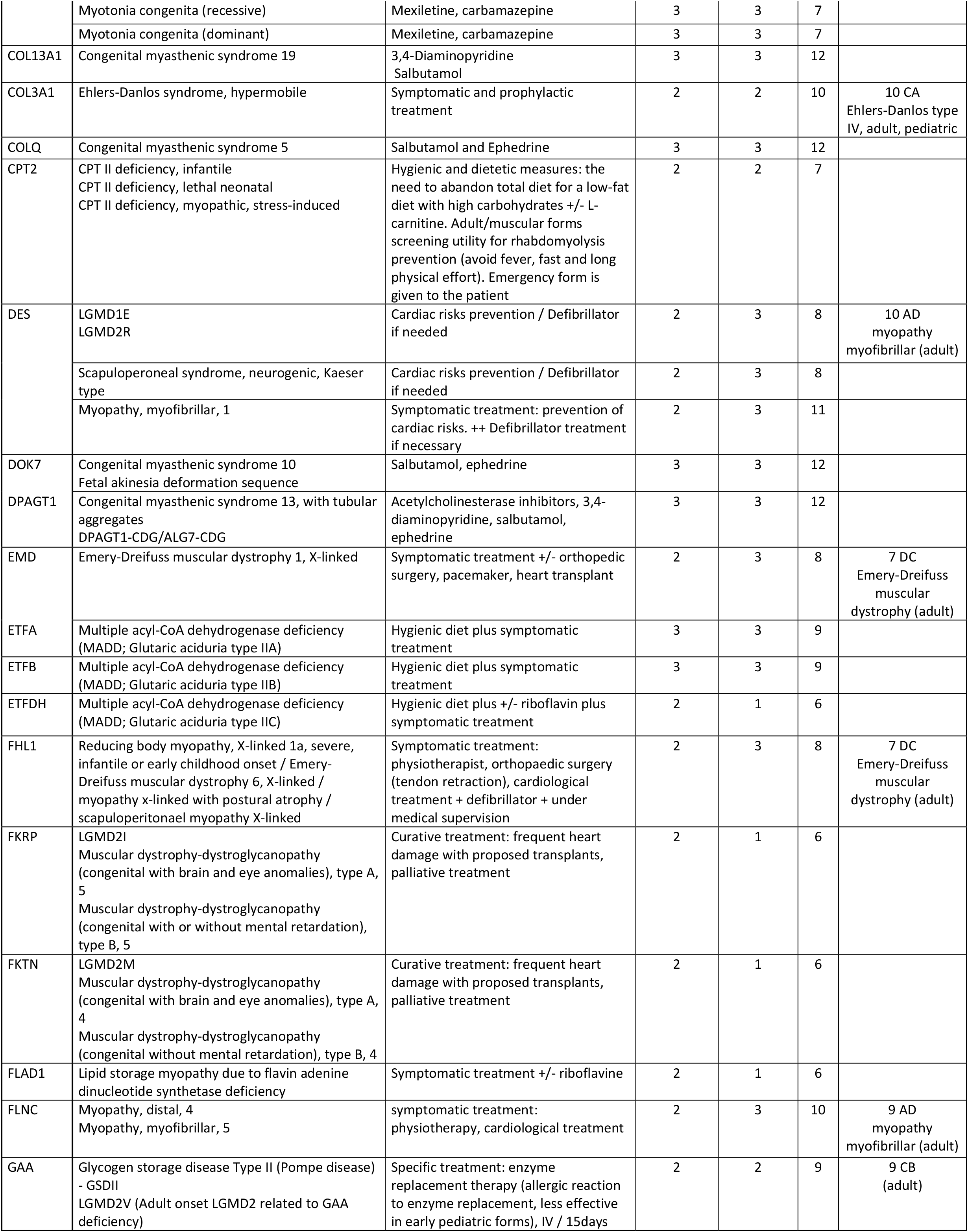
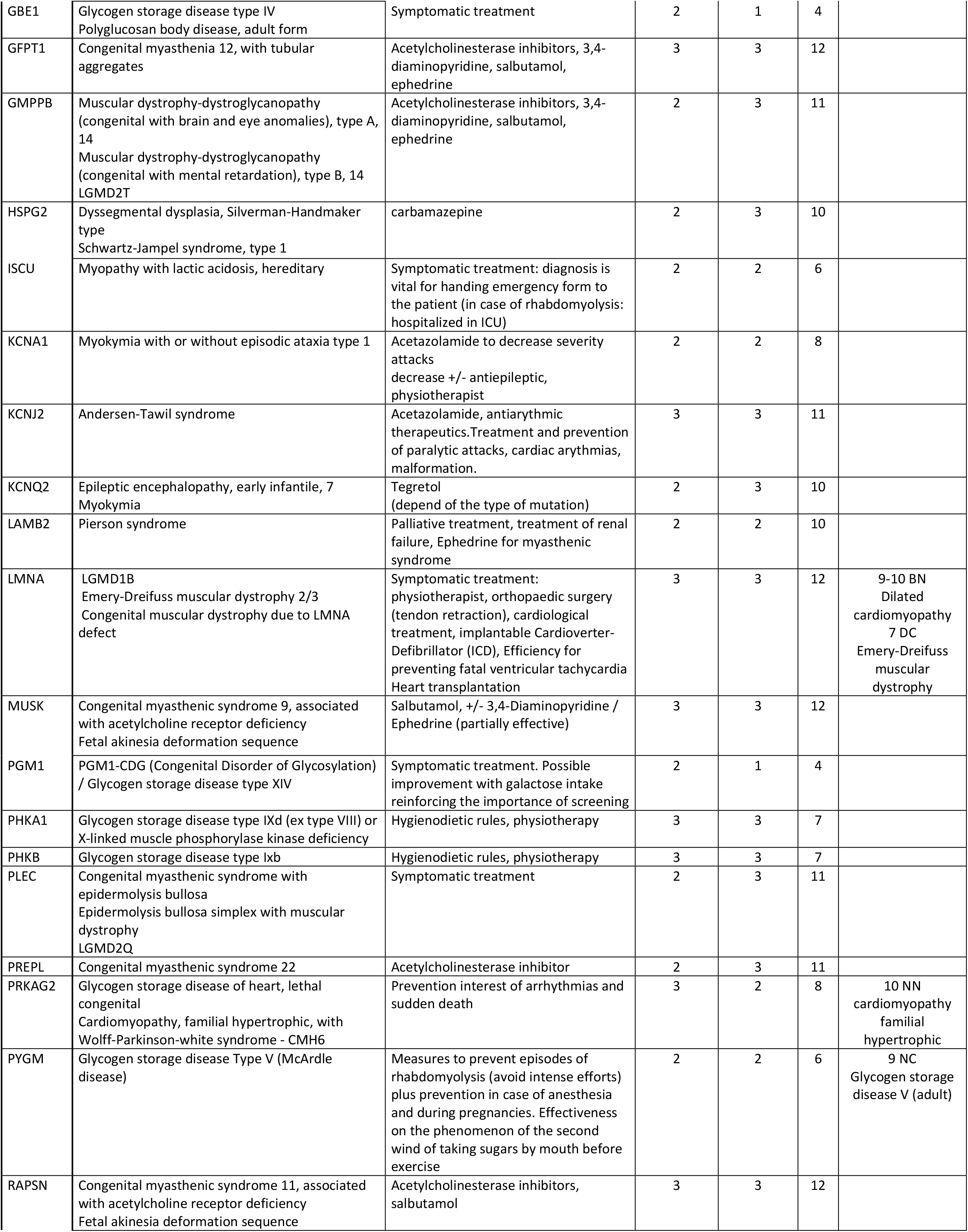
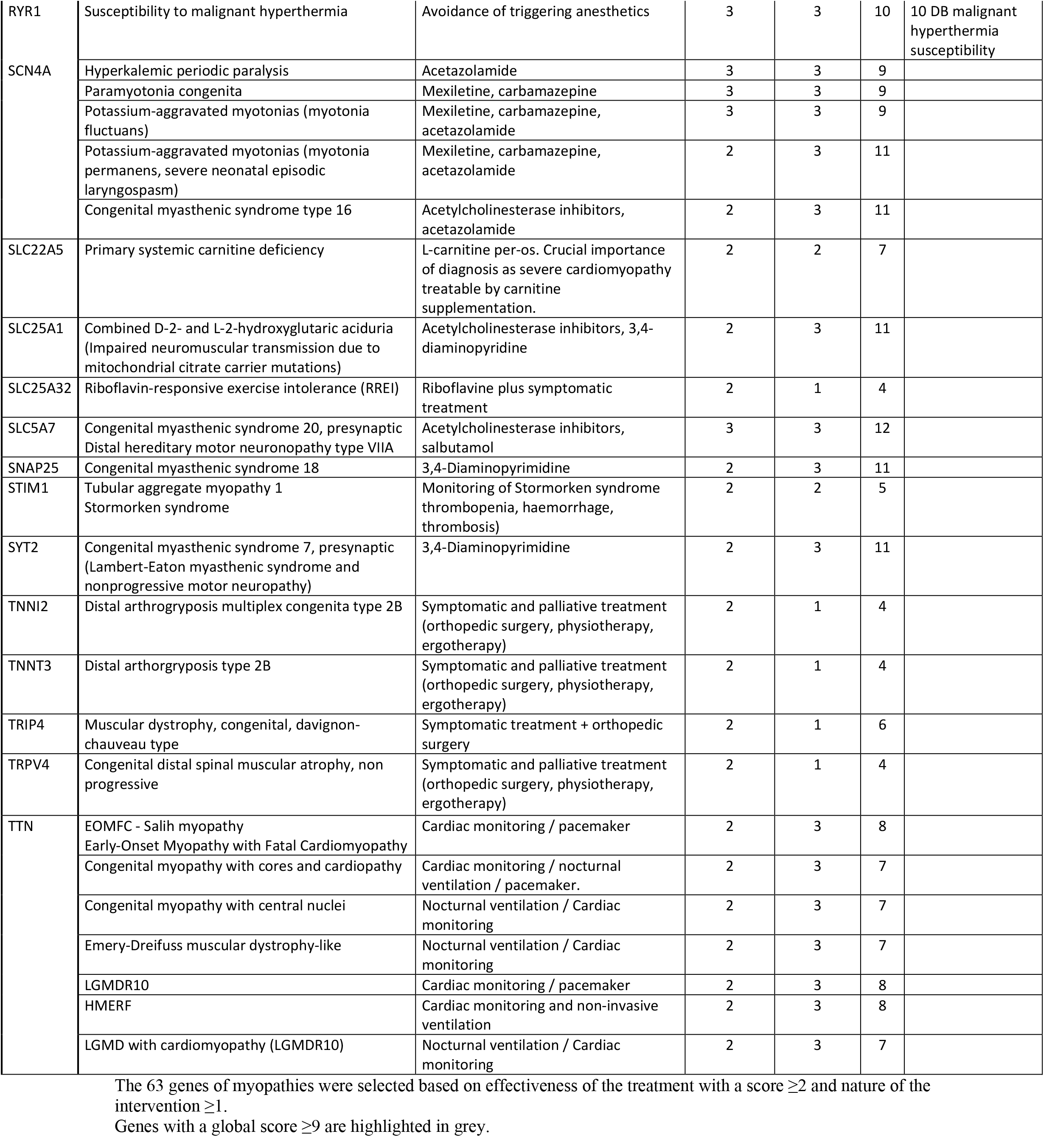
The 63 myopathy genes with a medical value for patient care.

We also evaluated the distribution of actionability scores within the 43 pairs genes/disorders (36 myopathy genes) with higher actionability scores (≥ 9) (fig 1c), of which only 8 had already been scored by ClinGen (Table 1). For the severity indicator: 77% had very high mortality rates, 14% had moderate severity and 9% low severity. For the disease penetrance indicator: the information was available for 95% of the genes and mostly with full penetrance (81%). The effectiveness of intervention was moderate (score 2) or high (score 3) for all genes. The nature of intervention was or moderate or low risk (scores 2 and 3) for 97% of genes. We then compared these actionability scores with those of ClinGen AWG genes with a total score ≥ 9 (111 genes corresponding to 78 disorders and 125 outcome-intervention pairs) (fig 1d). We found that the distribution of scores was different between both groups concerning the outcome-related domains (severity and likelihood), with higher scores for myopathy genes (p<0.05). Results for both intervention-related indicators are similar in the two categories of genes.

## DISCUSSION

We applied here, in an exhaustive way for the first time in the field of myopathies, the ClinGen AWG scoring methodological approach.

We encountered a lack of information on the penetrance data of myopathy genes in the literature (for 74% of genes), which was not the case for genes from the ClinGen AWG list genes (fig 1e)^4^. This lack of information on penetrance lead to a “loss” of 1 to 3 final scoring points, that could account for the lower final average actionability score of myopathy genes (score 4-5) compared to the genes scored by the ClinGen AWG (9-10). Noteworthy an important proportion of myopathies is of autosomal recessive inheritance, with expected complete penetrance. The use of the data collected by the French National Rare Diseases Data Bank (BAMARA) will possibly make it easier in the upcoming years to access data on penetrance for rare myopathies.

Thanks to our scoring work, we have identified 63 myopathy genes (corresponding to 78 disorders) that could be actionable on their management (intervention moderately of highly effective, without a high risk), including 43 genes/disorders with actionability scores ≥ 9. However, only 12 of these myopathy genes were previously scored by ClinGen. Of the 31 genes not scored by ClinGen AWG, 21 are part of congenital myasthenic syndromes (Table 1), for which there are specific treatments, reflecting the value of this scoring work ^7^.

Some of the 63 myopathy genes/disorders have a lower score of actionability (<9) but are considered to have a medical value for patient care. The *CLCN1* gene is a good example in the recessive or dominant myotonia congenita (Table 1). The low global score (7) is due to the modest morbidity (severity scored as 1), but the treatment is highly effective (scored as 3) and acceptable (scored as 3). For some genes associated with several diseases, the increase in the global score for actionability is due to an increase in the severity score. For example, the *SCN4A* gene is associated with an actionability score of 9 in the hyperkalemic periodic paralysis (severity score at 1), compared to a score of 11 in the Congenital myasthenic syndrome type 16 (severity score at 3) ^8^. Our results will allow to integrate the notion of actionable genes into strategic choices for molecular diagnostic strategies and management of patients with genetic myopathies. We recommend that these 63 genes be analyzed as a priority in the sequencing of gene panels, exomes or genomes, in order to offer a rapid diagnosis to patients and optimize the patient care (treatabolome database in preparation). Furthermore, these genes (at least the 36 with global score ≥ 9) should be considered for possible addition to the ACMG actionable gene list, in order to assess them as secondary data of medical value for patient care. Of course, patients’ phenotyping remains essential, and the absence of a mutation in this panel of genes does not exclude an intronic mutation in one of these genes. *DMD, DM1* and *DM2* genes were not the list of Filnemus genes analyzed by NGS.^5^ However, they should also be considered actionable, given their important consequences on patient management, as well as the *SMN1* gene.

The clinical actionability of genes must be considered as an evolving concept, in relation to progresses in therapeutic approaches, or diagnosis. In recent years new therapeutic options such as gene therapies have been developed and offer hope for patients with myopathies. It will be important to take these new therapeutic approaches into account in the coming years as several additional myopathic genes will then be considered actionable.

## Data Availability

All data produced in the present work are contained in the manuscript

## Abbreviations

ACMG: American College of Medical Genetics and Genomics
ClinGen: the Clinical Genome Resource
FILNEMUS: the French Network for Rare Neuromuscular Diseases

## Acknowledgments

We sincerely thank all the participants for collaborating work. This study was supported by the Filière nationale des Maladies Rares Neuromusculaires FILNEMUS. We thank Corinne Thèze for her help.

