## supplementary table 1 for "Objective evaluation of clinical actionability for genes involved in myopathies: 63 genes with a medical value for patient care"

| Gene | Disease | Severity | Severity detail | Penetrance | Intervention considered | Effectiveness | Nature of intervention | Knowledge base | Total score | pubmed | Score Clingen Officiel (Total score + Knowledge base) |
|---|---|---|---|---|---|---|---|---|---|---|---|
| ABHD5 | Triglyceride storage disease with impaired long-chain fatty acid oxidation (Chanarin-Dorfman syndrome) | 1 | Ichthyosis at birth, hepatomegalia, cataract, ataxia, deafness, mild muscular weakness, mild intellectual disability | 0 | Symptomatic treatment | 0 | 0 | E | 1 | 20691590<br>18769256 | |
| ACAD9 | Mitochondrial complex I deficiency due to ACAD9 deficiency | 2 | Childhood / adult: neurogical dysfunction, hepatic impairment, hypertrophic cardiomyopathy, exercise intolerance | 0 | Riboflavine | 2 | 2 | A | 6 | 22499348<br>30025539 | |
| ACADVL | Acyl-CoA dehydrogenase (very long chain) deficiency (VLCAD deficiency) | 3 | Neonatal form / late stage, severe form / mild, lethal in few months with metabolic acidosis, severe hypoketotic hypoglycemia, cardiomyopathy, hepatomegaly. Severe form, penetrance considered complete | 3 | Hygienic and dietetic measures +/- triheptanoin acid /Medium cahin triglyceride / N-acetylcystein<br>Prevention of rhabdomyolysis | 2 | 1 | C | 9 | 20301763<br>10077518<br>9973285<br>30993714<br>30477112 | |
| ACTA1 | Nemaline Myopathy 3- Actin congenital myopathy with cores<br>Actin congenital myopathy with excess of thin myofilaments<br>Congenital myopathy with fiber-type disproportion 1<br>Nemaline Myopathy 3 | 3 | Lethal respiratory insufficiency in the very early, neonatal form | 3 | Symptomatic treatment : physiotherapy, orthopaedic scoliosis surgery, breathing assistance | 1 | 1 | C | 8 | 31228046 | |
| ADAMTS2 | Ehlers-Danlos syndrome dermatosparaxis | 1 | Skin and joint hyperlaxity, umbilical and inguinal hernia, tissue fragility, dermatosparaxis, haematomas | 0 | Symptomatic treatment | 1 | 2 | E | 4 | 28306229 | |
| ADCY6 | Lethal congenital contracture syndrome 8 | 3 | Neonatal plus death in the neonatal period, decrease in active fetal movements, joint contractures, severe motor paralysis | 0 | Palliative treatment | 0 | 0 | E | 3 | 24319099 | |
| ADGRG6 | Lethal congenital contracture syndrome 9 | 3 | Lethal fetal form, Intrauterine Growth Retardation with pterygium, severe arthrogryposis, anasarca, hydramnios | 0 | Symptomatic and palliative treatment | 0 | 0 | E | 3 | 26004201 | |
| ADSSL1 | Myopathy, distal, 5 | 1 | Age of onset in adolescence, weakness and slowly progressive muscular atrophy predominant in the lower limbs | 0 | Symtomatic treatment | 1 | 2 | E | 4 | 26506222 | |
| AGL | Glycogen storage disease type IIIa - GSD IIIa<br>Glycogen storage disease type IIIb - GSD IIIb<br>Glycogen storage disease type IIIc - GSD IIIc<br>Glycogen storage disease type IIId - GSD IIId | 2 | Beginning in childhood, severe muscle weakness, hepatomegaly, hypertrophic cardiomyopathy (HCM) +/- heart failure and sudden death (rare), hepatomegaly +/- cirrhosis, stunded growth, hypotonia, hypoglycaemia-related convulsions | 0 | Hygienic and dietetic measures with specific diet<br>Vital disease screening because healthy measures can prevent hypoglycaemia and cardiac complication | 2 | 2 | C | 6 | 27106217<br>28888851 | |
| AGRN | Congenital myasthenic syndrome 8, with pre- and postsynaptic defects | 3 | Improvement of muscle strength, prevention of myasthenic decompensation | 3 | Salbutamol, ephedrine | 3 | 3 | B | 12 | 19631309<br>22205389 | |
| ALG14 | Congenital myasthenic syndrome 15, without tubular aggregates | 3 | Improvement of muscle strength, prevention of myasthenic decompensation | 3 | Acetylcholinesterase inhibitors, 3,4-diaminopyridine | 0 | 0 | C | 6 | 23404334<br>30808424 | |
| ALG2 | Congenital myasthenic syndrome 14, with tubular aggregates | 3 | Improvement of muscle strength, prevention of myasthenic decompensation | 3 | Acetylcholinesterase inhibitor (pyridostigmine) | 0 | 0 | C | 6 | 23404334<br>30808424 | |
| AMPD1 | Myopathy due to myoadenylate deaminase deficiency | 1 | Child / adult, asymptomatic / effort intolerance : cramp, tiredness | 0 | Symptomatic treatment / D-ribose | 2 | 1 | E | 4 | 28751290<br>12117480 | |
| ANO5 | LGMD2L<br>Miyoshi muscular dystrophy 3 (Early onset calf distal myopathy) | 1 | Adulthood, proximal muscle weakness / progressive distal damage, sometimes asymetrical, damage to the posterior quarters of the legs and thighs, variable expressiveness according to gender. Elevated CK > 1000 UI/L. Penetrating supposedly complete but variable expressivity | 3 | Symptomatic treatment | 1 | 2 | c | 7 | 20096397<br>23670307 | |
| ASCC1 | Spinal muscular atrophy with congenital bone fractures 2 | 3 | Neonatal, severe hypotonia in utero, bone fractures, arthrogrypose, difficulty repsiratory and food --> death 1st month/ years of life | 0 | Palliative treatment | 0 | 0 | E | 3 | 26924529<br>28218388<br>30327447 | |
| ATP2A1 | Brody myopathy | 1 | Beginning in childhood, muscle cramps and stiffness during exercise or in the cold, reversible in a few seconds, mainly upper and lower limb and head | 3 | Symptomatic treatment (physiotherapy, hot beths, avoidance of the cold) | 0 | 0 | B | 4 | 8841193<br>26248958 | |
| B3GALNT2 | Muscular dystrophy-dystroglycanopathy (congenital with brain and eye anomalies), type A, 11 | 2 | Variable symptomatology (early mucle weakness, intellectual disability, reduced life expectancy) | 0 | Palliative treatment / symptomatic | 1 | 2 | E | 5 | 23453667<br>24084573 | |
| B4GAT1<br>[B3GNT1] | Muscular dystrophy-dystroglycanopathy (congenital with brain and eye anomalies), type A, 13 | 3 | Possible early death, severe symptomatology: generalised muscular weakness, cerebral malformation, intellectual disability, cardiopathy | 0 | Palliative treatment | 1 | 2 | E | 6 | 23359570<br>23877401 | |
| BAG3 | Myofibrillar myopathy 6 | 3 | Early infancy, rapid development, generalised muscle weakness, respiratory failure in adolescence, cardiomyopathy +/- transplantation | 3 | Symptomatic treatment: Implantable Cardioverter-Defibrillator (ICD)/ ventilation | 2 | 3 | D | 11 | 26342832<br>30023292<br>25313375 | 9 AD<br>myopathy myofibrillar (adult) |
| BICD2 | Spinal muscular atrophy, lower extremity-predominant, 2 | 3 | Neonatal, multiple arthrogryposis, severe hypotonia, respiratory failure, early childhood death or joint deformation, muscle weakness, anterior horn motor neuron involvement, slowly progressive | 0 | Symptomatic and palliative treatment (orthopedic surgery, physiotherapy) | 0 | 0 | B | 3 | 23664116<br>23664119<br>23664120 | |
| BIN1 | Congenital myopathy<br>Centronuclear myopathy 2 | 2 | Variable hypotonia, muscular weakness, variable severitu, scoliosis, +/- severe respiratory disorders, +/- cardiomyopathy, good prognosis if no heart or respiratory attack | 0 | Symptomatic treatment (physiotherapy, orthopaedic surgery (scoliose)) | 1 | 2 | D | 5 | 25260562<br>17676042 | |
| BVES<br>[POPDC1] | LGMD2X | 2 | From adolescence onwards, progressive muscle weakness, arrhythmia and conduction disorder, syncopes | 0 | Symptomatic treatment | 1 | 2 | E | 5 | 26642364 | |
| CACNA1S | Congenital myopathy | 3 | Forms of congenital myopathy are very severe at birth | 0 | Symptomatic treatment | 1 | 1 | E | 5 | 8004673<br>28012042<br>26865514 | |
| | Susceptibility to malignant hyperthermia | 2 | Morbidity from malignant hyperthermia event | 2 | Avoidance of triggering anesthetics | 3 | 3 | | 10 | 16917943<br>16163667<br>19825159 | 10 DB<br>malignant hyperthermia susceptibility (adult) |
| | Hypokalemic periodic paralysis, type 1 (mutational hotspots in exons 4, 11, 21 and 30) | 1 | Periodic hypokalemic paralysis variable duration, beginning around 20 years +/- vacuolar myopathy --> permanent motor deficit at 40-50 years. Moderate congenital myopathy | 2 | Acétazolamide<br>Treatment and prevention of paralytic attacks | 3 | 3 | A | 9 | 8004673<br>7987325 | |
| CAPN3 | LGMD2A | 1 | Onset in adulthood, slight and progressive muscle weakness | 3 | Symptomatic treatment | 1 | 2 | C | 7 | 15725583<br>9771675 | |
| CASQ1 | Tubular aggregate myopathy (vacuolar myopathy with CASQ1 aggregates) | 1 | Begins at teen / adult age, muscle cramps + weakness, stable +/- asymptomatic | 2 | Symptomatic treatment | 0 | 0 | E | 3 | 26136523<br>25116801<br>30258016 | |
| CAV3 | LGMD1C<br>Distal myopathy, Tateyama type<br>Rippling muscle disease | 2 | Variable age of onset, muscle weakness and atrophy, Isolated elevated CK<br>Hypertrophic ou dilated cardiomyopathy<br>Long QT syndrome | 3 | Symptomatic treatment +/- orthopaedic surgery | 1 | 1 | C | 7 | 21496630<br>15580566<br>14981167<br>11431690<br>22581547 | |
| CCDC78 | Congenital myopathy (centronuclear myopathy 4) | 1 | Neonatal hypotonia, distal muscle weakness > proximal, significant fatigability, no cardiac/respiratory impairment, mild cognitive impairment | 0 | Symptomatic treatment | 0 | 0 | E | 1 | 22818856 | |
| CFL2 | Nemaline myopathy 7 | 1 | Muscular weakness, hypotonia, discrete respiratory impairment, slowly progressive | 0 | Symptomatic treatment | 1 | 2 | E | 4 | 22560515<br>17160903 | |
| CHAT | Congenital myasthenic syndrome 6 (presynaptic) | 3 | Improvement of muscle strength, prevention of myasthenic decompensation | 3 | Acetylcholinesterase inhibitors, 3,4-diaminopyridine, salbutamol, ephedrine | 3 | 2 | B | 11 | 26080897<br>30808424 | |
| CHKB | Congenital muscular dystrophy, megaconial type | 3 | Lethal cardiomyopathy | 0 | Symptomatic treatment plus vitamins | 1 | 1 | E | 5 | 29067961 | |
| CHRNA1 | Congenital myasthenic syndrome 1A<br>Congenital myasthenic syndrome 1B<br>Multiple pterygium syndrome, lethal type | 3 | Improvement of muscle strength, prevention of myasthenic decompensation | 3 | Fluoxetine, quinidine | 2 | 3 | B | 11 | 18252226<br>18179903<br>30808424<br>25888793 | |
| CHRNB1 | Congenital myasthenic syndrome 2C, associated with acetylcholine receptor deficiency<br>Congenital myasthenic syndrome 2A + Fetal akinesia deformation sequence | 3 | Improvement of muscle strength, prevention of myasthenic decompensation<br>Lethal fetal form, Intrauterine Growth Retardation with pterygium, severe arthrogryposis, anasarca, hydramnios | 3 | Fluoxetine, quinidine | 2 | 3 | B | 11 | 27364156<br>30808424 | |
| CHRND | Congenital myasthenic syndrome 3A<br>Congenital myasthenic syndrome 3B<br>Congenital myasthenic syndrome 3C<br>Multiple pterygium syndrome, lethal type | 3 | Improvement of muscle strength, prevention of myasthenic decompensation<br>lethal fetal form, Intrauterine Growth Retardation with pterygium, severe arthrogryposis, anasarca, hydramnios | 3 | Fluoxetine, quinidine | 2 | 3 | B | 11 | 18252226<br>18179903<br>30808424 | |
| CHRNE | Congenital myasthenic syndrome 4A, slow-channel | 3 | Improvement of muscle strength, prevention of myasthenic decompensatio | 3 | Fluoxetine, quinidine | 2 | 3 | B | 11 | 30808424<br>8357190<br>7531341<br>8957026 | |
| | Congenital myasthenic syndrome 4B, fast-channel | 3 | Improvement of muscle strength, prevention of myasthenic decompensation | 3 | Acetylcholinesterase inhibitors, salbutamol, ephedrine, 3,4-diaminopyridine | 3 | 3 | B | 12 | 30808424<br>8357190<br>7531341<br>8957026 | |
| | Congenital myasthenic syndrome 4C, associated with acetylcholine receptor deficiency | 3 | Improvement of muscle strength, prevention of myasthenic decompensation | 3 | Acetylcholinesterase inhibitors, 3,4-diaminopyridine, salbutamol, ephedrine | 3 | 3 | A | 12 | 30808424<br>8357190<br>7531341<br>8957026<br>11030414 | |
| CHRNG | Multiple pterygium syndrome, lethal type<br>Non lethal multiple pterygium syndrome (Escobar syndrome) | 3 | Lethal fetal form, intrauterine growth retardation (IUGR) with pterygium, severe arthrogryposis, anasarca, hydramnios)<br>(neonatal, respiratory insufficiency +/-, multiple pterygium, arthrogryposis | 3 | No effective treatment, sometimes try a symptomatic anticholinesterase-type treatment for fatigability | 0 | 0 | C | 6 | 16826531<br>18252226<br>30808424<br>18179903 | |
| CLCN1 | Thomsen Myotonia congenita<br>Becker congenita Myotonia | 1 | Myotonia, hypertrophy, difficulty in muscle relaxation, improvement with effort and worse with cold weather, variable severity | 2 | Mexiletine (side effect including abdominal pain) / lamotrigine / Ranolazine | 2 | 2 | A | 7 | 13797441<br>7981750 | |
| | Myotonia congenita (recessive) | 1 | Prevention of stiffness | 0 | Mexiletine, carbamazepine | 3 | 3 | B | 7 | 13797441<br>7981750 | |
| | Myotonia congenita (dominant) | 1 | Prevention of stiffness | 0 | Mexiletine, carbamazepine | 3 | 3 | C | 7 | 13797441<br>7981750 | |
| CNTN1 | Congenital myopathy (Compton-North) | 3 | Neonatal hypotonia, lack of spontaneous movement, muscle weakness respiratory failure, childhood death) | 0 | Palliative treatment | 0 | 1 | E | 4 | 19026398 | |
| CNTNAP1 | Lethal congenital contracture syndrome 7 | 3 | Lethal fetal form, Intrauterine Growth Retardation with pterygium, severe arthrogryposis, anasarca, hydramnios | 0 | Symptomatic and palliative treatment | 0 | 0 | E | 3 | 24319099 | |
| COL12A1 | Bethlem myopathy<br>Ullrich congenital muscular dystrophy<br>UCMD | 1 | Slowly progressive muscle weakness | 0 | Symptomatic treatment (physiotherapy, orthopaedic surgery (achilles tendon)) | 1 | 1 | C | 3 | 27348394<br>29342313 | |
| COL13A1 | Congenital myasthenic syndrome 19 | 3 | Improvement of muscle strength, prevention of myasthenic decompensation | 3 | 3,4-Diaminopyridine<br>Salbutamol | 3 | 3 | B | 12 | 26626625<br>30808424 | |
| COL1A1 | Ehlers-Danlos syndrome, classic<br>Ehlers-Danlos syndrome arthrochalasique | 1 | Joint hyperlaxity, +/- hips dislocation, hyperextensibility + skin fragility, muscular hypotonia | 0 | Symptomatic teatment (painkiller, arthrodesis, physiotherapy) | 1 | 2 | C | 4 | 28192633<br>27011056<br>28306229 | |
| COL1A2 | Ehlers-Danlos syndrome arthrochalasique | 1 | Joint hyperlaxity, +/- hips dislocation, hyperextensibility + skin fragility, muscular hypotonia | 0 | Symptomatic treatment (painkiller, arthrodesis, physiotherapy) | 1 | 2 | C | 4 | 28192633<br>28306229 | |
| COL3A1 | Ehlers-Danlos syndrome, hypermobile | 3 | Mostly : vascular complication of medium and large arteries, risk of death<br>More rarely : joint hyperlaxity, hyperextensibility + skin fragility, muscular hypotonia | 3 | Symptomatic and prophylactic treatment | 2 | 2 | C | 10 | 28192633<br>28306229 | 10 CA<br>Ehlers-Danlos type IV<br>adult (in released)<br>pediatric (in preparation) |
| COL5A1 | Ehlers-Danlos syndrome, classic | 1 | Hyperextensibility + skin fragility, atrophied scar, joint hyperlaxity +/- hips dislocation, muscular hypotonia | 0 | Symptomatic treatment (painkiller, arthrodesis, physiotherapy) | 1 | 3 | C | 5 | 28192633<br>23587214<br>27011056<br>28306229 | 8 ND / 5 DC<br>Ehlers-Danlos syndrome type 1 and 2 (adult) |
| COL5A2 | Ehlers-Danlos syndrome, classic | 1 | Hyperextensibility + skin fragility, atrophied scar, joint hyperlaxity +/- hips dislocation, muscular hypotonia | 0 | Symptomatic treatment (painkiller, arthrodesis, physiotherapy) | 1 | 3 | C | 5 | 28192633<br>23587214<br>27011056<br>28306229 | 8 ND / 5 DC<br>Ehlers-Danlos syndrome type 1 and 2 (adult) |
| COL5A3 | COL5A- / COL5A2-like phenotype | 1 | Hyperextensibility + skin fragility, atrophied scar, joint hyperlaxity +/- hips dislocation, muscular hypotonia +/- skin tumor | 0 | Symptomatic treatment (painkiller, arthrodesis, physiotherapy) | 1 | 3 | E | 5 | 10722718<br>26910848 | 8 ND / 5 DC<br>Ehlers-Danlos syndrome type 1 and 2 (adult) |

| | | | | | | | | | | | |
|---|---|---|---|---|---|---|---|---|---|---|---|
| COL6A1 | Bethlem myopathy/ Ullrich congenital muscular dystrophy | 2 | Slowly progressive proximal muscle weakness mixed with proximal disc retractions and hyperlaxity (stiff rachis). Ullrich congenital muscular dystrophy (UCMD): severe neonatal form characterised by respiratory insufficiency and severe disability. Bethlem myopathy : more moedrate form with loss of gait or worsening of gait at the age of 50, restrictive respiratory impairment is more moderate and rarely requires ventilation, unlike Ullrich's disease | 0 | Symptomatic treatment (physiotherapy, orthopaedic surgery (achilles tendon)) | 1 | 1 | C | 3 | 24443028<br>24938411<br>25204870<br>21496625 | |
| COL6A2 | Bethlem myopathy/ Ullrich congenital / Congenital myosclerosis | 2 | Slowly progressive proximal muscle weakness mixed with proximal disc retractions and hyperlaxity (stiff rachis). Ullrich congenital muscular dystrophy (UCMD): severe neonatal form characterised by respiratory insufficiency and severe disability. Bethlem myopathy : more moedrate form with loss of gait or worsening of gait at the age of 50, restrictive respiratory impairment is more moderate and rarely requires ventilation, unlike Ullrich's disease | 0 | Symptomatic treatment (physiotherapy, orthopaedic surgery (achilles tendon)) | 1 | 1 | C | 3 | 25204870<br>29406609<br>21496625 | |
| COL6A3 | Bethlem myopathy/ Ullrich congenital / Congenital myosclerosis | 2 | Slowly progressive proximal muscle weakness mixed with proximal disc retractions and hyperlaxity (stiff rachis). Ullrich congenital muscular dystrophy (UCMD): severe neonatal form characterised by respiratory insufficiency and severe disability. Bethlem myopathy : more moedrate form with loss of gait or worsening of gait at the age of 50, restrictive respiratory impairment is more moderate and rarely requires ventilation, unlike Ullrich's disease. Recessive forms of the Ullrich type are serious | 0 | Symptomatic treatment (physiotherapy, orthopaedic surgery (achilles tendon)) | 1 | 1 | C | 3 | 25204870<br>29406609<br>21496625 | |
| COL6A6 | COL6-like phenotype | 1 | Slowly progressive muscle weakness | 0 | Symptomatic treatment (physiotherapy, orthopaedic surgery (achilles tendon)) | 1 | 1 | E | 3 | 20882040<br>26247046 | |
| COLQ | Congenital myasthenic syndrome 5 | 3 | Improvement of muscle strength, prevention of myasthenic decompensation | 3 | Salbutamol<br>Ephedrine | 3 | 3 | A | 12 | 9758617<br>9689136<br>30808424 | |
| CPT2 | CPT II deficiency, infantile<br>CPT II deficiency, lethal neonatal<br>CPT II deficiency, myopathic, stress-induced | 3 | Begins a few months of life, severe fasting intolerance, hypoglycaemia with hypoacetonaemia --> coma / convulsions, +/- myopathy and cardiomyopathy with fatal paroxysmal arrhythmia | 0 | Hygienic and dietetic measures : the need to abandon total diet for a low fat diet with high carbohydrates +/- L-carnitine. Adult/muscular forms screening utility for rhabdomyolysis prevention (avoid fever, fast and long physical effort). Emergency form is given to the patient | 2 | 2 | C | 7 | 10607472<br>20691590 | |
| CRYAB | Myopathy, myofibrillar, fatal infantile hypertrophy, alpha-B crystallin-related | 3 | Begins a few weeks of life, rapidly progressing muscular rigidity, lethal respiratory failure before 3 years, cardiomyopathy, cataract, less severe adult AD formation | 0 | Palliative treatment | 0 | 0 | E | 3 | 20171888<br>27389816<br>9731540<br>14681890 | |
| DAG1 | Muscular dystrophy-dystroglycanopathy (congenital with brain and eye anomalies), type A, 9<br>LGMD2P (Recessive LGMD with primary alphaDG defect) | 2 | LGMD2P and MDD : slowly progressive muscle weakness, +/- intellectual disability (ID) | 0 | Symptomatic treatment, palliative treatment for Sd Walker-Warburg | 1 | 2 | C | 5 | 14678799<br>25503980 | |
| DES | LGMD1E<br>LGMD2R | 3 | Onset in adulthood, more or less severe muscle weakness, more or less heart disease with possible sudden death, more or less respiratory failure | 0 | Cardiac risks prevention / Defibrillator if needed | 2 | 3 | D | 8 | 10717012 | 10 AD<br>myopathy myofibrillar (adult) |
| | Scapuloperoneal syndrome, neurogenic, Kaeser type | 3 | Early adulthood, muscle weakness +/- severe, +/- heart disease with possible sudden death, +/- respiratory failure | 0 | Cardiac risks prevention / Defibrillator if needed | 2 | 3 | D | 8 | 10717012<br>25313375 | |
| | Myopathy, myofibrillar, 1 | 3 | Onset in adulthood, more or less severe muscle weakness, more or less heart disease with possible sudden death, more or less respiratory failure | 3 | Symptomatic treatment: prevention of cardiac risks.<br>++ Defibrillator treatment if necessary | 2 | 3 | D | 11 | 9697706<br>10545598<br>10717012<br>20718792 | |
| DPM3 | DPM3-CDG (Congenital Disorder of Glycosylation) | 2 | Progressive muscle weakness, dilated cardiomyopathy with or without rhythm disturbance | 0 | Symptomatic treatment | 1 | 2 | C | 5 | 19576565<br>28803818<br>31266720 | |
| DNAJB6 | LGMD1D | 1 | Variable age of onset, slowly progressive muscle weakness, distal and proximal forms | 0 | Symptomatic treatment | 1 | 2 | C | 4 | 22366786<br>26205529 | |
| DNM2 | Centronuclear myopathy 1<br>Charcot-Marie-Tooth disease, axonal type 2M<br>Charcot-Marie-Tooth disease, dominant intermediate B<br>Lethal congenital contracture syndrome 5 | 3 | Early childhood / adult, muscle weakness / +/- slowly progressive peripheral neuropathy, AR form haemorrhagic fetal akinesia, severe hypotonia, respiratory failure, joint contraction, lethal | 3 | Symptomatic treatment (physiotherapy, orthopaedic surgery) / palliative treatment for fetal akinesia | 1 | 1 | C | 8 | 23092955<br>30208955<br>27861221<br>19502294<br>22091729 | |
| DOK7 | Congenital myasthenic syndrome 10<br>Fetal akinesia deformation sequence | 3 | Improvement of muscle strength, prevention of myasthenic decompensation<br>lethal fetal form, Intrauterine Growth Retardation with pterygium, severe arthrogryposis, anasarca, hydramnios | 3 | Salbutamol, ephedrine | 3 | 3 | A | 12 | 19261599<br>30588388 | |
| DOLK | DOLK-CDG (Congenital Disorder of Glycosylation) | 2 | Dilated cardiomyopathy: death in childhood, hypotonia, muscle weakness, multi-system involvement, ichthyosis, epilepsy, progressive microcephaly | 0 | Symptomatic treatment | 1 | 2 | C | 5 | 22242004<br>17273964 | |
| DPAGT1 | Congenital myasthenic syndrome 13, with tubular aggregates<br>DPAGT1-CDG/ALG7-CDG | 3 | Improvement of muscle strength, prevention of myasthenic decompensation | 3 | Acetylcholinesterase inhibitors, 3,4-diaminopyridine, salbutamol, ephedrine | 3 | 3 | B | 12 | 22742743<br>23447650<br>30808424 | |
| DPM1 | DPM1-CDG (Congenital Disorder of Glycosylation) | 2 | Psychomotor delay, convulsion, hypotonia, dysmorphia microcephaly | 0 | Symptomatic treatment | 1 | 2 | D | 5 | 10642602<br>10642597<br>23856421<br>29079546 | |
| DPM2 | DPM2-CDG (Congenital Disorder of Glycosylation) | 2 | Delayed growth and development, osteopenia, hypotonia, liver dysfunction, severe epilepsy with of without early death | 0 | Symptomatic treatment | 1 | 2 | D | 5 | 23109149<br>19901254 | |
| DPM3 | DPM3-CDG (Congenital Disorder of Glycosylation) | 2 | progressive muscle weakness, dilated cardiomyopathy with or without rhythm disturbance | 0 | symptomatic treatment | 1 | 2 | C | 5 | 19576565<br>28803818<br>31266720 | |
| DYNC1H1 | Spinal muscular atrophy, lower extremity-predominant 1 | 1 | Early childhood, weakness and muscular atrophy of the lower limbs, stable/ very slowly progressive | 0 | Symptomatic treatment | 1 | 1 | B | 3 | 22459677<br>25609763 | |
| DYSF | LGMD2B<br>Miyoshi muscular dystrophy 1<br>Distal myopathy with anterior tibial onset | 2 | Begins in adulthood, progressive muscle weakness, severe myopathy, very marked handicap | 0 | Symptomatic treatment | 1 | 2 | C | 5 | 17698709<br>31043956<br>31019989 | |
| ECEL1 | Distal arthrogryposis type 5D | 1 | Neonatal contractures of feet and hands, facial dysmorphia | 0 | Symptomatic treatment | 1 | 1 | B | 3 | 23236030<br>23261301 | |
| EMD | Emery-Dreifuss muscular dystrophy 1, X-linked | 3 | Onset in childhood, weakness plus muscle atrophy, tendon retractions, scoliosis, cardiomyopathy, rhythm disorder, conduction disorder : sudden death, respiratory failure | 0 | Symptomatic treatment +/- orthopedic surgery, pacemaker, heart transplant | 2 | 3 | C | 8 | 20301609<br>23622360 | 7 DC<br>Emery-Dreifuss muscular dystrophy (adult) |
| ENO3 | Glycogen storage disease XIII GSD13 | 1 | Adult, exercise intolerance, myalgia, McArdle-Like penotype. Very rare disease | 0 | Symptomatic treatment | 0 | 0 | E | 1 | 25267339<br>11506403<br>30397902 | |
| ERBB3 | Lethal congenital contractural syndrome 2 | 3 | Neonatal with death, arthrogrypose pultiple, craniofacial abnormality, anterior horn cell degeneration | 0 | Symptomatic and palliative treatment | 0 | 0 | E | 3 | 17701904 | |
| ETFA | Multiple acyl-CoA dehydrogenase deficiency (MADD; Glutaric aciduria type IIA) | 3 | Neonatal form / late form, severe / less severe, lethal within a few months with metabolic acidosis, severe hypoketotic hypoblycaemia, cardiomyopathy, hepatomegaly. High diagnostic interest as the disease is mostly treatable by riboflavin supplementation plus or minus carnitine. | 0 | Hygienic diet plus symptomatic treatment | 3 | 3 | C | 9 | 12815589<br>25200064 | |
| ETFB | Multiple acyl-CoA dehydrogenase deficiency (MADD; Glutaric aciduria type IIB) | 3 | Neonatal form / late form, severe / less severe, lethal within a few months with metabolic acidosis, severe hypoketotic hypoblycaemia, cardiomyopathy, hepatomegaly. High diagnostic interest as the disease is mostly treatable by riboflavin supplementation plus or minus carnitine. | 0 | Hygienic diet plus symptomatic treatment | 3 | 3 | B | 9 | 12815589<br>25200064 | |
| ETFDH | Multiple acyl-CoA dehydrogenase deficiency (MADD; Glutaric aciduria type IIC) | 3 | Neonatal form / late form, severe / less severe, lethal within a few months with metabolic acidosis, severe hypoketotic hypoblycaemia, cardiomyopathy, hepatomegaly | 0 | Hygienic diet plus +/- riboflavin plus symptomatic treatment | 2 | 1 | E | 6 | 12815589 | |
| FBLN5 | Cutis laxa, autosomal dominant 2<br>Cutis laxa, autosomal recessive, type IA<br>Neuropathy, hereditary, with or without age-related macular degeneration | 3 | Loss of skin elacsticity, from birth onwards, lethal cardiovascular and respiratory damage in the autosomal recessive (AR) form before 20 years | 0 | Symptomatic treatment | 1 | 1 | C | 5 | 28383366 | |
| FBN2 | Congenital contractural arachnodactyly | 3 | Early neonatal, Marfan-like syndrome, death in ploymalformative form with heart/ digestive involvement, severe scoloise, flexion joint contraction, arachnodactylia, muscle hypoplasia | 3 | Symptomatic and palliative treatment (orthopedic surgery, physiotherapy) | 1 | 1 | B | 8 | 7493032<br>11754102<br>18767143<br>19006240 | |
| FHL1 | Reducing body myopathy, X-linked 1a, severe, infantile or early childhood onset / Emery-Dreifuss muscular dystrophy 6, X-linked / myopathy x-linked with postural atrophy / scapuloperitonael myopathy X-linked | 3 | Respiratory failure, arrhythmia, conduction disorder, hypertrophic cardiomyopathy (HCM) then dilated cardiomyopathy (DCM) : sudden death | 0 | Symptomatic treatment: physiotherapist, orthopaedic surgery (tendon retraction), cardiological treatment + defibrillator + under medical supervision | 2 | 3 | C | 8 | 29735270<br>25313375 | 7 DC<br>Emery-Dreifuss muscular dystrophy (adult) |
| FKBP14 | Ehlers-Danlos syndrome with progressive kyphoscoliosis, myopathy, and hearing loss | 1 | Congenital muscle hypotonia, congenital or early kyphoscoliosis, joint hyperlaxity / subluxations, skin fragility, blue sclera, deafness | 0 | Symptomatic treatment | 1 | 1 | C | 3 | 28306229 | |
| FKRP | LGMD2I<br>Muscular dystrophy-dystroglycanopathy (congenital with brain and eye anomalies), type A, 5<br>Muscular dystrophy-dystroglycanopathy (congenital with or without mental retardation), type B, 5 | 3 | Severe if Walker Warburg Syndrome, Isolated elevated CK, dilated cardiomyopathy, LGMD2I: slowly progressive muscle weakness, dilated cardiomyopathy, respiratory muscle damage | 0 | Curative treatment : frequent heart damage with proposed transplants, palliative treatment | 2 | 1 | C | 6 | 17878207<br>15883334 | |
| FKTN | LGMD2M<br>Muscular dystrophy-dystroglycanopathy (congenital with brain and eye anomalies), type A, 4<br>Muscular dystrophy-dystroglycanopathy (congenital without mental retardation), type B, 4 | 3 | Possible early death, severe symptomatology: generalized muscle weakness, brain malformation, intelectual disability (ID), heart disease. Rare but existing mild forms. Isolated elevated CK | 0 | Curative treatment : frequent heart damage with proposed transplants, palliative treatment | 2 | 1 | C | 6 | 17878207<br>19179078 | |
| FLAD1 | Lipid storage myopathy due to flavin adenine dinucleotide synthetase deficiency | 3 | Childhood: heart and respiratory failure : early death, adult: muscle weakness | 0 | Symptomatic treatment +/- riboflavine | 2 | 1 | E | 6 | 27259049<br>28433476<br>30680745 | |
| FLNC | Myopathy, distal, 4<br>Myopathy, myofibrillar, 5 | 2 | Progressive muscle weakness with respiratory weakness and cardiomyopathy, conduction disorder and rhythm disorder | 3 | symptomatic treatment: physiotherapy, cardiological treatment | 2 | 3 | E | 10 | 15824355<br>21620354<br>25313375 | 9 AD<br>myopathy myofibrillar (adult) |
| GAA | Glycogen storage disease Type II (Pompe disease) - GSDII<br>LGMD2V (Adult onset LGMD2 related to GAA deficiency) | 3 | Differentiate between infantile (infantile-onset Pompe disease IOPD) and adult (late-onset Pompe disease LOPD) forms. Major benefit of screening for infantile forms because treatment with enzyme replacement therapy (ERT) should be started early. But there is also a great interest in detecting adult forms, even those that are not very severe, because respiratory complications can be prevented | 3 | Specific treatment: enzyme replacement therapy (allergic reaction to enzyme replacement, less effective in early paediatric forms)<br>IV / 15days | 2 | 2 | A | 9 | 28477382<br>16702877<br>8558570<br>22253258 | 9 CB<br>(adult) |
| GBE1 | Glycogen storage disease type IV<br>Polyglucosan body disease, adult form | 1 | Lethal fetal form / congenital, hepatomegaly plus hypotonia plus delayed development plus or minus cirrhosis with portal hypertension plus or minus cardiomyopathy and respiratory failure. Polyglucosan: neurogenic bladder in adults, loss of sensation in extremities. Mainly liver disease with rare early neuromuscular forms (arthrogryposis) | 0 | Symptomatic treatment | 2 | 1 | C | 4 | 25728520<br>27546458<br>30397902 | |
| GFPT1 | Congenital myasthenia 12, with tubular aggregates | 3 | Improvement of muscle strength, prevention of myasthenic decompensation | 3 | Acetylcholinesterase inhibitors, 3,4-diaminopyridine, salbutamol, ephedrine | 3 | 3 | A | 12 | 21310273<br>21975507<br>30808424<br>30808424 | |
| GLE1 | Lethal congenital contracture syndrome 1<br>Arthrogryposis with Anterior Horn Cell Disease | 3 | Neonatal death before 32 Weeks of gestation +++, total fetal akinesia, anasarch, micrognathia, pulmonary hypoplasia, multiple acticular contractures | 0 | Symptomatic and palliative treatment | 0 | 0 | E | 3 | 18204449<br>27684565 | |
| GMPPB | Muscular dystrophy-dystroglycanopathy (congenital with brain and eye anomalies), type A, 14<br>Muscular dystrophy-dystroglycanopathy (congenital with mental retardation), type B, 14<br>LGMD2T | 3 | Improvement of muscle strength, prevention of myasthenic decompensation | 3 | Acetylcholinesterase inhibitors, 3,4-diaminopyridine, salbutamol, ephedrine | 2 | 3 | B | 11 | 23768512<br>26133662 | |
| GNE | Distal myopathy with rimmed vacuoles (Nonaka) and Hereditary inclusion body myopathy | 1 | Slowly progressive muscle weakness | 0 | Symtomatic treatment and diet | 1 | 2 | C | 4 | 30842975<br>29720219 | |
| GOLGA2 | Developmental delay, seizures, progressive microcephaly, and muscular dystrophy<br>Mucopolysaccharidosis type 3 | 2 | Microcephaly, hypotonia, febrile infantile spasms, developmental delay | 0 | Symptomatic treatment | 0 | 0 | E | 2 | 26742501 | |

| | | | | | | | | | | | |
|---|---|---|---|---|---|---|---|---|---|---|---|
| GYG1 | Glycogen storage disease type XV<br>Polyglucosan body myopathy 2 | 2 | Variable onset, distal muscle involvement more or less scapuloperoneal, more or less cardiac involvement (hypertrophic cardiomyopathy CMH) and ventricular arrhythmia, slowly progressive. | 0 | Symptomatic treatment | 1 | 1 | E | 3 | 20357282<br>29264399 | |
| GYS1 | Glycogen storage disease, type 0 | 3 | Early childhood hypertrophic cardiomyopathy (CMH) plus arrhythmia : sudden death, muscle fatigue plus pain, syncope during physical activity | 0 | Symptomatic treatment | 1 | 2 | E | 6 | 18358695<br>19699667 | |
| HACD1<br>[PTPLA] | Congenital myopathy | 2 | Variable hypotonia, variable muscle weakness severity, scoliosis, respiratory problems +/- severe, +/- cardiomyopathy, good prognosis if not cardiac/respiratory involvement | 0 | Symptomatic treatment ( orthopedic surgery, physiotherapy ) | 1 | 1 | E | 4 | 23933735 | |
| HNRNPA1 | Inclusion body myopathy with early-onset Paget disease without frontotemporal dementia 3 | 3 | Adult, multi-system degenerative disorder, muscle weakness, Paget's disease, frontotemporal dementia, repiratory/ heart failure : death | 0 | Symptomatic treatment | 1 | 2 | C | 6 | 23455423<br>27066560 | |
| HNRNPDL | LGMD1G | 1 | adult onset, mild and progressive muscle weakness | 0 | symptomatic treatment | 1 | 2 | E | 4 | 31267206<br>24647604<br>15367920 | |
| HSPG2 | Dyssegmental dysplasia, Silverman-Handmaker type<br>Schwartz-Jampel syndrome, type 1 | 2 | bone dysplasia, more or less neonatal form of dwarfism if onset early after birth, facial dysmorphia, joint contractures, more or less urogenital / cardiovascular abnormality, myotonia | 3 | carbamazepine | 2 | 3 | A | 10 | 11101850 | |
| INPP5K | Muscular dystrophy, congenital, with cataracts and intellectual disability | 2 | Muscle damage plus intelectual disability (ID) plus cataract plus respiratory failure | 0 | symptomatic treatment + hormonology if hypogonadism | 1 | 1 | E | 4 | 28190456<br>28190459 | |
| ISCU | Myopathy with lactic acidosis, hereditary | 2 | Severe exercise intolerance in childhood: heart palpitations, lactic acidosis, fatigue, muscle weakness plus or minus extensive muscle paralysis and circulatory shock: possible death. | 0 | Symptomatic treatment: diagnosis is vital for handing emergency form to the patient (in case of rhabdomyolysis: hospitalized in ICU) | 2 | 2 | C | 6 | 18304497<br>18296749<br>28007899 | |
| ISPD | Muscular dystrophy-dystroglycanopathy (congenital with brain and eye anomalies), type A, 7<br>LGMD2U (Limb-Girdle, Muscular dystrophy related to ISPD) | 3 | Possible early death, severe symptomatology: generalized muscle weakness, brain malformation, intelectual disability (ID), heart disease | 0 | Palliative treatment | 1 | 2 | E | 6 | 22522421 | |
| ITGA7 | Congenital muscular dystrophy due to ITGA7 deficiency | 1 | Hypotonia, neonatal, dyspnea due to respiratory muscle weakness , scoliosis muscle weakness, predominantly proximal and atrophy Delayed motor development, cognitive impairment (in 1 of 3 patients) | 0 | Symptomatic treatment | 1 | 1 | E | 3 | 19330236<br>19260934 | |
| KBTBD13 | Nemaline myopathy 6 | 1 | Muscle weakness of the hands and feet, discreet respiratory damage, slowly progressive | 0 | Symptomatic treatment | 1 | 2 | C | 4 | 21109227 | |
| KCNA1 | Myokymia with or without episodic ataxia type 1 | 1 | In childhood, spastic contractions of skeletal muscles, ataxia, more or less epilepsy | 3 | Acetazolamide to decrease severity attacks decrease +/- antiepileptic, physiotherapist | 2 | 2 | E | 8 | 7842011<br>19307729 | |
| KCNJ2 | Andersen-Tawil syndrome | 2 | Periodic paralysis, QT interval prolongation plus ventricular arrhythmias: sudden death, scoliosis | 3 | Acetazolamide, antiarythmic therapeutics.Treatment and prevention of paralytic attacks, cardiac arythmias, malformation. | 3 | 3 | B | 11 | 12148092 | |
| KCNQ2 | Epileptic encephalopathy, early infantile, 7<br>Myokymia | 2 | Benign familial neonatal epilepsy (BFNE) // Neonatal epileptic encephalopathy (NEE) | 3 | Tegretol<br>(depend of the type of mutation) | 2 | 3 | A | 10 | 11572947<br>17872363<br>30771507 | |
| KLHL40 | Nemaline myopathy 8 | 3 | Neonatal, severe with death in childhood, respiratory failure, heart disease, arthrogryposis, muscle contractions | 0 | Palliative treatment | 0 | 0 | C | 3 | 27528495<br>23746549 | |
| KLHL41 | Nemaline myopathy 9 | 3 | Neonatal onset, hypotonia, retraction, severe respiratory failure | 0 | Symptomatic treatment / palliative treatment | 0 | 0 | C | 3 | 24268659 | |
| KLHL9 | Distal myopathy | 1 | Begins around 10-20 years old, weakness + slowly progressive distal muscle atrophy of the limbs, loss of tenderness at the extremities | 0 | Symptomatic treatment | 1 | 2 | E | 4 | 20554658 | |
| KY | Congenital myopathy with core targetoid Myopathy, myofibrillar, 7 | 1 | Begins in childhood, progressive muscle weakness, muscle contracture | 0 | Symptomatic treatment ( orthopaedic surgery, physiotherapy ) | 1 | 1 | E | 3 | 27484770<br>27485408 | |
| LAMA2 | Muscular dystrophy, congenital merosin-deficient - MDC1A | 3 | Respiratory failure and arrhythmia | 0 | Symptomatic treatment (orthopaedic surgery for scoliosis, noninvasive ventilation (VNI), physiotherapy) | 1 | 1 | C | 5 | 18700894<br>16216942 | |
| LAMB2 | Pierson syndrome | 3 | Improvement of muscle strength, prevention of myasthenic decompensation, treatment of kidney disease | 3 | Palliative treatment, treatment of renal failure, Ephedrine for myasthenic syndrome | 2 | 2 | B | 10 | 19251977<br>30808424 | |
| LAMP2 | Danon disease | 3 | Severe cardiomyopathy with frequent mental retardation and sometimes retinopathy | 0 | Symptomatic treatment +/- transplant | 1 | 1 | E | 5 | 26748608<br>29753918<br>30857840 | |
| LARGE1<br>[LARGE] | Muscular dystrophy-dystroglycanopathy (congenital with brain and eye anomalies), type A, 6<br>Muscular dystrophy-dystroglycanopathy (congenital with mental retardation), type B, 6 | 2 | Variable symptomatology (early muscle weakness, reduced life expectancy) or severe in the fetus | 0 | Symptomatic treatment / palliative treatment | 1 | 2 | E | 5 | 19067344<br>17436019<br>24709677 | |
| LDB3<br>[ZASP] | Late onset distal myopathy (Markesbery-Griggs)<br>Myopathy, myofibrillar, 4 | 2 | Variable age of onset, heart disease, progressive distal / proximal muscle weakness | 0 | Symptomatic treatment | 1 | 1 | E | 4 | 26342832<br>27389816<br>15668942<br>27546599<br>23263837 | |
| LDHA | Glycogen storage disease XI - GSD11 | 1 | Exercise intolerance: fatigue, pain and muscle cramps<br>McArdle-Like phenotype. Very rare disease | 0 | Symptomatic treatment | 0 | 0 | E | 1 | 29198466<br>7603529 | |
| LIMS2 | LGMD2W (Limb girdle muscular dystrophy with severe cardiomyopathy and triangular tongue) | 3 | Onset in childhood, weakness plus progressive and severe muscle atrophy, severe dilated cardiomyopathy | 0 | Symptomatic treatment | 1 | 2 | E | 6 | 25589244 | |
| LMNA | LGMD1B<br>Emery-Dreifuss muscular dystrophy 2/3<br>Congenital muscular dystrophy due to LMNA defect | 3 | Respiratory failure, arrhythmia, conduction disorder, dilated cardiomyopathy (DCM) : sudden death | 3 | Symptomatic treatment: physiotherapist, orthopaedic surgery (tendon retraction), cardiological treatment<br>Implantable Cardioverter-Defibrillator (ICD)<br>Efficiency for preventing fatal ventricumar tachycardia<br>Heart transplantation | 3 | 3 | B | 12 | 10080180<br>10814726<br>15668447<br>17377071<br>16407522<br>10739764<br>18551513<br>31155932<br>27938454 | 9-10 BN<br>Dilated cardiomyopathy (adult)<br><br>7 DC<br>Emery-Dreifuss muscular dystrophy (adult) |
| LMOD3 | Nemaline myopathy 10 | 3 | Neonatal onset, hypotonia, retraction, severe respiratory failure, broad phenotypic spectrum | 0 | Symptomatic and palliative treatment | 1 | 1 | C | 5 | 25250574 | |
| LPIN1 | Acute recurrent myoglobinuria | 3 | Childhood, frequent and severe rhabdomyolysis, hypertonia, muscle stiffness, renal failure. Potentially very serious disease with many deaths of children in episodes of rhabdomyolysis related to rhythm disorders. | 0 | Symptomatic treatment: Glucose and IV hydratation, monitoring of vital functions. Very high importance of diagnosis to give emergency cards and to hospitalise urgently at the first symptoms of rhabdomyolysis. | 1 | 2 | E | 5 | 20583302 | |
| LRP4 | Congenital myasthenic syndrome 17 | 3 | Improvement of muscle strength, prevention of myasthenic decompensation | 3 | Salbutamol, ephedrine | 0 | 0 | C | 6 | 24234652<br>30808424 | |
| MAGEL2 | Schaaf-Yang syndrome | 2 | Arthrogryposis, scoliosis, obesity, spychomotor dvpt delay, Intelectual Disability, autism traits, neonatal hypotonia | 0 | Symptomatic treatment (orthopedic surgery, physiotherapy) | 1 | 1 | B | 4 | 26365340<br>28281571<br>29359444 | |
| MAP3K20<br>[ZAK] | Centronuclear myopathy 6 with fiber-type disproportion | 1 | Begins at birth / first years, hypotonia + generalized muscle weakness, joint contractures, scoliosis, stable/ slowly progressive. | 0 | Symptomatic treatment | 0 | 0 | E | 1 | 27816943 | |
| MATR3 | Amyotrophic lateral sclerosis 21<br>Vocal cord and pharyngeal distal myopathy | 1 | Muscle weakness in the hands and feet, swallowing and voice problems | 0 | Symptomatic treatment: physiotherapy and speech therapy | 1 | 2 | E | 4 | 25154462<br>23842731<br>19344878<br>28872913 | |
| MEGF10 | Congenital myopathy (early-onset myopathy with areflexia, respiratory distress and dysphagia) | 2 | Neonatal onset with severe hypotonia, low generalized msucular +/- severe, respiratory failure +/-severe --> childhood death respiratory arrest, scoliosis, cleft palate | 0 | Symptomatic treatment | 1 | 1 | E | 4 | 22101682<br>29128256 | |
| MTM1 | Congenital myopathy<br>(X-linked myotubular myopathy) | 3 | Classic form: neonatal onset, lethal respiratory failure, muscle weakness, hypotonia, deaths in early life most of the time. Rarer form: onset in children/adults, male and female, muscle weakness of varying severity plus or minus respiratory problems, good prognosis if no respiratory involvement. | 3 | Palliative treatment | 0 | 1 | A | 7 | 8640223<br>24070817<br>28685322<br>22968136<br>30902907<br>28237839 | |
| MUSK | Congenital myasthenic syndrome 9, associated with acetylcholine receptor deficiency<br>Fetal akinesia deformation sequence | 3 | Improvement of muscle strength, prevention of myasthenic decompensation | 3 | Salbutamol, +/- 3,4-Diaminopyridine / Ephedrine (partially effective) | 3 | 3 | A | 12 | 25537362<br>25612909<br>30808424 | |
| MYBPC1 | Distal arthrogryposis type 1B<br>Lethal congenital contracture syndrome 4 | 3 | Lethal respiratory failure in the first hours of life, multiple and severe joint contractions, muscle atrophy. Campodactylie, varus equine feet, varaible and slowly progressive phenotype. | 0 | Symptomatic and palliative treatment | 1 | 1 | E | 5 | 22610851 | |
| MYBPC3 | Congenital myopathy | 3 | Neonatal myopathy + lethal cardiomyopathy | 0 | Palliative treatment | 0 | 0 | E | 3 | 19858127 | |
| MYH2 | Congenital myopathy (proximal myopathy and ophthalmoplegia)<br>Distal arthrogryposis | 2 | Muscle weakness, +/- joint contractures at birth, variable severity, stable / very slowly progressive, moderate phenotype in severity. | 0 | Symptomatic treatment | 1 | 1 | E | 4 | 24193343<br>23489661 | |
| MYH3 | Distal arthrogryposis, type 2A / type 2B / type 8<br>Freeman-Sheldon (DA2A)<br>Sheldon-Hall (DA2B)<br>Multiple pterygium syndrome (DA8) | 2 | Distal forms of arthrogrypose, feet/ hands, +/- scoliosis, dysmorphia | 0 | Symptomatic treatment (orthopedic surgery, physiotherapy) | 1 | 1 | A | 4 | 16642020<br>25256237<br>25957469 | |
| MYH7 | Laing distal myopathy<br>Congenital myopathy (myosin storage myopathy)<br>Scapuloperoneal syndrome, myopathic type | 2 | Begins in childhood/ adult age, highly variable phenotype, more severe AR with heart disease +/- sudden death, slowly progressive muscle weakness and atrophy, scoliosis | 3 | Symptomatic treatment ( orthopedic surgery, physiotherapy, cardiology ) | 1 | 2 | B | 8 | 24664454<br>27387980 | |
| MYH8 | Trismus-pseudocamptodactyly syndrome | 1 | Trismus plus neonatal pseudocamptodactylia | 0 | Symptomatic treatment | 1 | 1 | B | 3 | 15282353<br>17041932 | |
| MYMK | Carey-Fineman-Ziter syndrome | 2 | Begins at birth, hypotonia, moebius sequence, Pierre-Robin sequence, severe scoliosis, stunting | 0 | Symptomatic treatment (orthopedic surgery, oto-rhino-laryngology, physiotherapy) | 1 | 1 | E | 4 | 28681861 | |
| MYO18B | Klippel-Feil syndrome 4 with myopathy and facial dysmorphism | 3 | Neonatal onset, central hypotonia, cardiomyopathy, more or less pulmonary arterial hypertension (PAH), hemodynamic instability, myopathy, facial dysmorphia, possible early death | 0 | Symptomatic and palliative treatment | 1 | 1 | E | 5 | 27858739<br>25748484 | |
| MYO9A | Congenital myasthenic syndrome, type not numbered yet<br>Presynaptic congenital myasthenic syndrome | 3 | Improvement of muscle strength, prevention of myasthenic decompensation | 3 | Acetylcholinesterase inhibitor +/- 3,4-Diaminopyridine | 0 | 0 | C | 6 | 27259756<br>30808424 | |
| MYOD1 | Fetal akinesia deformation sequence | 3 | Lethal fetal form, IUCN with pterygium, severe arthrogryposis, anasarque, hydramnios | 0 | Symptomatic and palliative treatment | 0 | 0 | E | 3 | 26733463 | |
| MYOT | LGMD1A<br>Myopathy, myofibrillar, 3<br>Myopathy, spheroid body | 1 | Early adulthood, muscle weakness +/- severe, +/- heart disease, +/- respiratory failure. Myotiline: distal myopathy often beginning around the 5th decade or more or less severe proximal forms. | 0 | Symtomatic treatment | 1 | 2 | D | 4 | 10958653<br>16380616 | |
| MYPN | Nemaline myopathy 11 | 1 | Muscle weakness, hypotonia, discrete respiratory impairment, slowly progressive | 0 | Symptomatic treatment | 1 | 2 | E | 4 | 28017374<br>28220527 | |
| NALCN | Congenital contractures of the limbs and face, hypotonia, and developmental delay<br>Infantile hypotonia, with psychomotor retardation and characteristic facies 1 | 2 | Neonatal onset, persistent severe hypotonia, global psychomotor retardation + intellectual impairment, dysmorphia, growth retardation, +/- congenital contractures. | 0 | Symptomatic treatment +/- acétazolamide | 1 | 1 | B | 4 | 25683120<br>25864427<br>24075186<br>27214504 | |
| NEB | Nemaline Myopathy 2 | 2 | Extended phenotypic spectrum | 0 | Symptomatic and palliative treatment | 1 | 2 | C | 5 | 12207937<br>25205138 | |
| ORAI1 | Tubular aggregate myopathy 2 | 1 | Begins in childhood/adolescence, diffuse muscle weakness, hypocalcemia, slowly progressive | 0 | Symptomatic treatment | 0 | 0 | E | 1 | 31448844<br>25227914 | |
| PFKM | Glycogen storage disease Type VII (Tarui) | 1 | Early childhood, muscle exercise intolerance, infantile form can be rapidly fatal, more or less dilated cardiomyopathy and breathing difficulty | 0 | Avoid physical exercise, no treatment | 0 | 0 | E | 1 | 747977610<br>24427140 | |
| PGAM2 | Glycogen storage disease X - GSD10 | 1 | Exercise-induced cramps<br>McArdle-Like phenotype. Very rare disease | 0 | Symptomatic treatment | 0 | 0 | E | 1 | 23169535<br>28779239 | |
| PGK1 | Phosphoglycerate kinase 1 deficiency | 1 | Chronic haemolytic anaemia, exercise intolerance, muscle weakness, cramps, myalgia, intelectual disability, more or less epilepsy, more or less ataxia, more or less trembling, neuromuscular signs like CMT | 0 | No treatment | 0 | 0 | E | 1 | 16740138<br>19157875<br>26883264<br>30887539 | |
| PGM1 | PGM1-CDG (Congenital Disorder of Glycosylation) /<br>- Glycogen storage disease type XIV | 2 | Neonatal, Pierre Robin sequence plus cleft palate, intermittent hypoglycaemia, exercise intolerance, chronic hepatitis, multi-systemic disease. Muscle involvement dominated by rhabdomyolysis with possible cardiac involvement | 0 | Symptomatic treatment. Possible improvement with galactose intake reinforcing the importance of screening | 2 | 1 | C | 4 | 24499211 | |

| | | | | | | | | | | | |
|---|---|---|---|---|---|---|---|---|---|---|---|
| PHKA1 | Glycogen storage disease type IXd (ex type VIII) or X-linked muscle phosphorylase kinase deficiency | 1 | Adolescent / adult: muscle damage with exercise intolerance, myalgia, cramps and more or less asymptomatic fatigue<br>McArdle-like pathology more benign than McArdle. | 0 | Hygienodietic rules, physiotherapy | 3 | 3 | C | 7 | 15637709<br>12825073<br>30659246 | |
| PHKB | Glycogen storage disease type Ixb | 1 | Child: muscle damage with exercise intolerance, myalgia, cramps and fatigue, more or less symptomatic hypotonia and liver damage, more or less symptomatic interventricular septal hypertrophy / adults are asymptomatic | 0 | Hygienodietic rules, physiotherapy | 3 | 3 | B | 7 | 9215682<br>30659246 | |
| PIEZO2 | Distal arthrogryposis type 3<br>Arthrogryposis, muscle weakness and scoliosis<br>Distal arthrogryposis type 5 | 2 | Reserved prognosis in some cases, neonatal, severe hypotonia, multiple foot malformation syndrome, hands +/- heart, cyphoscoliosis | 0 | Symptomatic treatment (orthopedic surgery, physiotherapy) | 1 | 1 | B | 4 | 23487782<br>27843126<br>27974811<br>24726473<br>25712306<br>30941898 | |
| PIP5K1C | Lethal congenital contractural syndrome 3 | 3 | Neonatal onset with death, multiple arthrogrypose, craniofacial abnormality, anterior horn cell degeneration | 0 | Symptomatic and palliative treatment | 0 | 0 | E | 3 | 17701898 | |
| PLEC | Congenital myasthenic syndrome with epidermolysis bullosa<br>Epidermolysis bullosa simplex with muscular dystrophy<br>LGMD2Q | 3 | Improvement of muscle strength, prevention of myasthenic decompensation, prevention and treatment of skin disease | 3 | Symptomatic treatment | 2 | 3 | C | 11 | 8696340<br>10446808 | |
| PLOD1 | Ehlers-Danlos syndrome cyphoscoliotic | 2 | Congenital muscle hypotonia, congenital or early kyphoscoliosis, joint hyperlaxity/subluxations, skin fragility, bluish sclera, eyeball fragility | 3 | Symptomatic treatment | 1 | 1 | C | 7 | 28306229 | |
| PNPLA2 | Neutral lipid storage disease with myopathy without ichthyosis | 2 | Adult, limb muscle weakness, slowly progressive, 50% cardiomyopathy, diabetes, fatty liver disease | 0 | No treatment | 0 | 0 | E | 2 | 18657972<br>21544567 | |
| PNPLA8 | Mitochondrial myopathy with lactic acidosis (MMLA) | 1 | From the first years of life proximate muscle weakness, dystonia, dysmetria, spasticity, partial complex epilepsy, progressive lactic acidosis | 0 | Symptomatic treatment | 0 | 0 | E | 1 | 25512002 | |
| POGLUT1 | LGMD2Z | 1 | Young adult, weakness plus slowly progressive muscle atrophy, one case with breathing difficulties | 0 | Symptomatic treatment | 1 | 2 | E | 4 | 27807076 | |
| POMGNT1 | LGMD2O<br>Muscular dystrophy-dystroglycanopathy (congenital with brain and eye anomalies), type A, 3<br>Muscular dystrophy-dystroglycanopathy (congenital with mental retardation), type B, 3 | 3 | Severe if Walker Warburg syndrome, other MDD depressive syndrome<br>LGMD2O: slowly progressive muscle weakness. | 0 | Symptomatic and palliative treatment | 1 | 2 | C | 6 | 15236414<br>19067344 | |
| POMGNT2 | Muscular dystrophy-dystroglycanopathy (congenital with brain and eye anomalies), type A, 8 | 3 | Severe if Walker Warburg syndrome, other MDD depressive syndrome<br>LGMD2O: slowly progressive muscle weakness | 0 | Symptomatic and palliative treatment | 1 | 2 | D | 6 | 19067344<br>15236414 | |
| POMK | Muscular dystrophy-dystroglycanopathy (congenital with brain and eye anomalies), type A, 12<br>Muscular dystrophy-dystroglycanopathy (limb-girdle), type C, 12 | 2 | Variable symptomatology (early muscle weakness, intelectual disability, microcephaly, reduced life expectancy) | 0 | Symptomatic and palliative treatment | 1 | 2 | E | 5 | 24556084<br>24925318 | |
| POMT1 | Muscular dystrophy-dystroglycanopathy (congenital with brain and eye anomalies), type A, 1<br>Muscular dystrophy-dystroglycanopathy (congenital with mental retardation), type B, 1<br>LGMD2K | 3 | Severe if Walker Warburg syndrome<br>LGMD2K: slowly progressive muscle weakness, intelectual disability, microcephaly) | 0 | Palliative treatment except for LGMD2I symptomatic treatment | 1 | 2 | C | 6 | 17878207<br>12369018 | |
| POMT2 | LGMD2N<br>Muscular dystrophy-dystroglycanopathy (congenital with brain and eye anomalies), type A, 2<br>Muscular dystrophy-dystroglycanopathy (congenital with mental retardation), type B, 2 | 2 | Slowly progressive muscle weakness, intelectual disability | 0 | Symptomatic treatment | 1 | 2 | C | 5 | 17878207<br>19138766 | |
| PREPL | Congenital myasthenic syndrome 22 | 3 | Improvement of muscle strength, prevention of myasthenic decompensation | 3 | Acetylcholinesterase inhibitor | 2 | 3 | C | 11 | 24610330<br>30808424 | |
| PRKAG2 | Glycogen storage disease of heart, lethal congenital<br>Cardiomyopathy, familial hypertrophic, with Wolff-Parkinson-white syndrome - CMH6 | 3 | Severe arrhythmogenic heart disease more or less lethal in the neonatal period, cardiac congestion: arrest, pulmonary oedema | 0 | Prevention interest of arrhythmias and sudden death | 3 | 2 | C | 8 | 28431061 | 10 NN<br>cardiomyopathy familial hypertrophic (adult) |
| PYGM | Glycogen storage disease Type V (McArdle disease) | 1 | Beginning in childhood, intolerance to effort: myalgias, cramps, fatigue and muscle weakness, resolving after a few minutes of rest, more or less if formed at birth: hypotonia and respiratory insufficiency | 0 | Measures to prevent episodes of rhabdomyolysis (avoid intense efforts) plus prevention in case of anesthesia and during pregnancies. Effectiveness on the phenomenon of the second wind of taking sugars by mouth before exercise | 2 | 2 | A | 6 | 21802952<br>29143597 | 9 NC<br>Glycogen storage disease V (adult) |
| PYROXD1 | Early-Onset myopathy with internalized nuclei and myofibrillar disorganization | 1 | Childhood, slowly progressive muscle weakness, muscle atrophy | 0 | Symptomatic treatment | 0 | 0 | E | 1 | 27745833<br>31455395<br>30515627 | |
| RAPSN | Congenital myasthenic syndrome 11, associated with acetylcholine receptor deficiency<br>Fetal akinesia deformation sequence | 3 | Improvement of muscle strength, prevention of myasthenic decompensation | 3 | Acetylcholinesterase inhibitors, salbutamol | 3 | 3 | A | 12 | 15328566<br>12730725<br>12796535<br>18252226<br>18179903<br>30808424 | |
| RBCK1 | Polyglucosan body myopathy 1 with or without immunodeficiency | 3 | Childhood, progressive proximate muscle weakness, severe lethal cardiomyopathy plus or minus transplantation plus or minus severe immunodeficiency and hyperinflamation. Most patients have severe heart disease, the cause of death. Crucial importance of diagnosis of this very rare disease | 0 | Symptomatic treatment +/- transplant | 1 | 1 | E | 5 | 23798481<br>23889995<br>25041762 | |
| RYR1 | Susceptibility to malignant hyperthermia | 2 | Morbidity from malignant hyperthermia event | 2 | Avoidance of triggering anesthetics | 3 | 3 | A | 10 | | 10 DB malignant hyperthermia susceptibility (adult) |
| | Central core disease<br>King-Denborough syndrome<br>Minicore myopathy with external ophthalmoplegia<br>Congenital neuromuscular disease with uniform type 1 fiber (RYR1-related congenital myopathy with fatigable weakness, responding to pyridostigimine) | 2 | Spinal rigidity, muscle weakness and atrophy, hypotonia, developmental delay, more or less malignant hyperthermia, stable and slowly progressive | 3 | Symptomatic treatment (orthopedic surgery for scoliosis, physiotherapy). Systematic screening of patients at risk of malignant hyperthermia and recommendations for anesthesia (against indication: depolarizing curares, halogenated anesthetics) | 1 | 1 | B | 7 | 30406384<br>28818389 | |
| SCN4A | Hyperkalemic periodic paralysis | 1 | Treatment and prevention of paralytic attacks | 2 | Acetazolamide | 3 | 3 | C | 9 | 26700687 | |
| | Paramyotonia congenita | 1 | Prevention of stiffness and weakness episodes | 2 | Mexiletine, carbamazepine | 3 | 3 | B | 9 | 26700687 | |
| | Potassium-aggravated myotonias (myotonia fluctuans) | 1 | Prevention of stiffness. | 2 | Mexiletine, carbamazepine, acetazolamide. | 3 | 3 | A | 9 | 26700687 | |
| | Potassium-aggravated myotonias (myotonia permanens, severe neonatal episodic laryngospasm) | 3 | Prevention of stiffness, treament and prevention of laryngospasm | 3 | Mexiletine, carbamazepine, acetazolamide | 2 | 3 | A | 11 | 26700687 | |
| | Congenital myasthenic syndrome type 16 | 3 | Improvement of muscle strength, prevention of myasthenic decompensation | 3 | Acetylcholinesterase inhibitors, acetazolamide | 2 | 3 | B | 11 | 26700687 | |
| SELENON [SEPN1] | Rigid spine muscular dystrophy 1 /<br>- Congenital myopathy with fiber-type disproportion | 2 | Minor to severe at birth muscle weakness, stable in 90% of cases, progressive repiratory failure, rigid spine | 0 | Symptomatic treatment (orthopedic surgery, physiotherapy) | 1 | 1 | E | 4 | 11528383<br>15122708<br>16365872<br>12192640<br>20937510<br>21670436 | |
| SGCA | LGMD2D | 2 | Onset in childhood, progressive muscle weakness, more or less cardiac involvement in 20% of cases: reduced life expectancy | 0 | Symptomatic treatment | 1 | 2 | C | 5 | 7663524<br>30838895 | |
| SGCB | LGMD2E | 2 | Onset in childhood, progressive muscle weakness, varying degrees of cardiac and respiratory impairment | 0 | Symptomatic treatment | 1 | 2 | C | 5 | 28284983<br>10662809<br>10678176 | |
| SGCD | LGMD2F | 2 | Variable age of onset, progressive muscle weakness, respiratory impairment and more or less severe cardiomyopathy with early death | 0 | Symptomatic treatment | 1 | 2 | C | 5 | 8841194<br>10662809 | |
| SGCG | LGMD2C | 2 | Onset in childhood, progressive muscle weakness, more or less cardiac and respiratory impairment: premature death | 0 | Symptomatic treatment | 1 | 2 | C | 5 | 22240777<br>31194043<br>18285821 | |
| SLC22A5 | Primary systemic carnitine deficiency | 3 | Early childhood/adult, progressive heart disease: heart failure / sudden death, muscle weakness, hypotonia, recurrent hypoglycemic / hypoketotic seizures more or less coma, lethargy. Crucial importance of diagnosis as severe heart disease treatable by carnitine supplementation. | 0 | L-carnitine per-os. Crucial importance of diagnosis as severe cardiomyopathy treatable by carnitine supplementation. | 2 | 2 | C | 7 | 10051646<br>11058897 | |
| SLC25A1 | Combined D-2- and L-2-hydroxyglutaric aciduria (Impaired neuromuscular transmission due to mitochondrial citrate carrier mutations) | 3 | Improvement of muscle strength, prevention of myasthenic decompensation | 3 | Acetylcholinesterase inhibitors, 3,4-diaminopyridine | 2 | 3 | C | 11 | 26870663<br>30808424 | |
| SLC25A20 | Carnitine-acylcarnitine translocase deficiency | 3 | Neonatal, severe hypoketotic hypoglycemia, hyperammonia, cardiomyopathy with or without arrhythmia, liver failure, muscle weakness, encephalopathy. | 0 | Hygienodietetic rules: avoid fasting, carbihydrates, medium chain triglycerides, unsaturated poly fatty acids, carnitine. | 1 | 1 | E | 5 | 15363639<br>15365988<br>30477112 | |
| SLC25A32 | Riboflavin-responsive exercise intolerance (RREI) | 1 | Childhood, exercise intolerance | 0 | Riboflavine plus symptomatic treatment | 2 | 1 | E | 4 | 26933868<br>28443623 | |
| SLC5A7 | Congenital myasthenic syndrome 20, presynaptic<br>Distal hereditary motor neuronopathy type VIIA | 3 | Improvement of muscle strength, prevention of myasthenic decompensation | 3 | Acetylcholinesterase inhibitors, salbutamol | 3 | 3 | B | 12 | 27569547<br>29088354<br>30808424 | |
| SNAP25 | Congenital myasthenic syndrome 18 | 3 | Improvement of muscle strength, prevention of myasthenic decompensation, management of intellectual disability and ataxia | 3 | 3,4-Diaminopyrimidine | 2 | 3 | C | 11 | 25792100<br>30808424 | |
| SPEG | Congenital myopathy (centronuclear myopathy 5) | 3 | Neonatal onset with severe hypotonia, low muscle +/- severe, respiratory failure and heart failure +/-severe --> childhood death | 0 | Symptomatic treatment | 1 | 2 | E | 6 | 25087613<br>28624463<br>29614691<br>30412272 | |
| SPTBN4 | Myopathy, congenital, with neuropathy and deafness | 2 | Begins in neonatal period, severe hypotonia, scoliosis, generalized muscle atrophy, central deafness, +/- epilepsy. | 0 | Symptomatic treatment | 0 | 0 | E | 2 | 28540413 | |
| SQSTM1 | Distal myopathy with rimmed vacuoles | 1 | aearly adulthood, muscle weakness and muscular atrophy | 0 | symtomatic treatment | 1 | 2 | E | 4 | 26208961<br>27594680 | |
| STAC3 | Congenital myopathy (Native American myopathy)<br>Baily-Bloch | 2 | Muscle weakness, arthrogrypose, cyphoscoliosis, cleft palate, palpebral ptosis, malignant hyperthermia from anesthesia | 0 | Symptomatic treatment ( orthopedic surgery, physiotherapy ) | 1 | 1 | C | 4 | 28777491<br>30168660 | |
| STIM1 | Tubular aggregate myopathy 1<br>Stormorken syndrome | 1 | Exercise cramps/ weakness, alcohol/drug induced myopathy, starts in childhood, ophthalmoparesis | 0 | Monitoring of Stormorken syndrome thrombopenia, haemorrhage, thrombosis) | 2 | 2 | C | 5 | 31448844<br>23332920<br>24570283 | |
| SYT2 | Congenital myasthenic syndrome 7, presynaptic (Lambert-Eaton myasthenic syndrome and nonprogressive motor neuropathy) | 3 | Improvement of muscle strength, prevention of myasthenic decompensation | 3 | 3,4-Diaminopyrimidine | 2 | 3 | C | 11 | 25192047<br>29874875<br>30808424 | |
| TCAP | LGMD2G | 2 | Weakness plus atrophy of proximal lower limb muscles +/- respiratory impairment and hypertrophic cardiomyopathy | 0 | Symptomatic treatment | 1 | 1 | E | 3 | 24843229<br>10655062<br>16352453 | |
| TIA1 | Welander distal myopathy | 1 | Begins around 40 years, slowly progressive muscle weakness and atrophy of the hands | 0 | Symptomatic treatment: physiotherapy | 1 | 2 | E | 4 | 23401021<br>23348830 | |
| TMEM5 | Muscular dystrophy-dystroglycanopathy (congenital with brain and eye anomalies), type A, 10 | 2 | Variable symptomatology (early muscle weakness, intelectual disability, reduced life expectancy) or very severe in fetuses | 0 | Symptomatic treatment or palliative treatment | 1 | 2 | E | 5 | 23217329 | |
| TNNI2 | Distal arthrogryposis multiplex congenita type 2B | 1 | From birth, joint contracture feet + hands, camptodactylia, dysmorphia | 0 | Symptomatic and palliative treatment (orthopedic surgery, physiotherapy, ergotherapy) | 2 | 1 | B | 4 | 12592607<br>16802141<br>27790376 | |
| TNNT1 | Nemaline myopathy 5 | 3 | Neonatal onset, hypotonia, retraction, severe respiratory failure, life expectancy of 2 years | 0 | Palliative treatment. | 0 | 0 | C | 3 | 29931346 | |
| TNNT3 | Distal arthorgryposis type 2B | 1 | From birth, joint contracture feet + hands, camptodactylia, dysmorphia | 0 | Symptomatic and palliative treatment (orthopedic surgery, physiotherapy, ergotherapy) | 2 | 1 | C | 4 | 12865991<br>25337069<br>21402185 | |
| TNPO3 | LGMD1F | 2 | Variable age onset, proximal and distal muscle weakness, respiratory failure | 0 | Symptomatic treatment | 1 | 2 | C | 5 | 23667635<br>23543484 | |
| TNXB | Ehlers-Danlos syndrome due to tenascin X deficiency | 1 | Hyperextensibility plus skin fragility, velvety skin, upapyratic scars, joint hyperlaxity, spontaneaous bruising | 0 | Symptomatic treatment | 1 | 2 | E | 4 | 28306229<br>27582382<br>23768946 | |

| | | | | | | | | | | |
|---|---|---|---|---|---|---|---|---|---|---|
| TOR1AIP1 | LGMD2Y | 3 | From 10-20 years, weakness plus slowly progressive muscle atrophy, stiff spine, retractions, limited respiratory function, mild cardiomyopathy more or less cardiac arrest | 0 | Symptomatic treatment | 1 | 2 | E | 6 | 24856141 |
| TPM2 | Nemaline Myopathy 4<br>CAP myopathy 2<br>Distal arthrogryposis multiplex congenita type 1<br>Distal arthrogryposis type 2B | 2 | Muscle weakness of the hands and feet, discreet respiratory damage, slowly progressive | 0 | Symptomatic treatment ( orthopedic surgery, physiotherapy), respiratory assistance | 1 | 1 | C | 4 | 24692096 |
| TPM3 | Nemaline Myopathy 1<br>Congenital myopathy with fiber-type disproportion<br>CAP myopathy 1 | 2 | Begins at birth/ 1st year, hypotonia, minor to severe generalized muscle weakness if AR, moderate to severe respiratory damage | 0 | Symptomatic treatment +/- respiratory assistance +/- palliative treatment | 1 | 2 | C | 4 | 24692096 |
| TRAPPC11 | LGMD2S | 1 | Slowly progressive muscle weakness | 0 | Symptomatic treatment + orthopedic surgery | 1 | 1 | E | 3 | 24843229<br>23830518 |
| TRIM32 | LGMD2H | 2 | Beginning 20-30 years, slowly progressive muscle weakness more or less respiratory | 0 | Symptomatic treatment | 1 | 1 | C | 4 | 30823891<br>11822024<br>25351777 |
| TRIP4 | Muscular dystrophy, congenital, davignon-chauveau type | 3 | Severe respiratory failure<br>frequent cardiac involvement | 0 | Symptomatic treatment + orthopedic surgery | 2 | 1 | E | 6 | 27008887<br>26924529 |
| TRPV4 | Congenital distal spinal muscular atrophy, non progressive | 1 | Neonatal onset, atrophy + stable and distal muscle weakness in the lower limbs, bending joint contractures in the lower limbs, bladder and intestinal dysfunction | 0 | Symptomatic and palliative treatment (orthopedic surgery, physiotherapy, ergotherapy) | 2 | 1 | B | 4 | 25900305<br>24789864 |
| TTN | EOMFC - Salih myopathy<br>Early-Onset Myopathy with Fatal Cardiomyopathy | 3 | Congenital or infantile muscle weakness with axial and distal joint contractures cardiomyopathy adolescence lethal | 0 | Cardiac monitoring / pacemaker | 2 | 3 | A | 8 | 24980681 |
| | Congenital myopathy with cores and cardiopathy | 2 | Congenital or infantile muscle weakness, cardiomyopathy | 0 | Cardiac monitoring / nocturnal ventilation / pacemaker. | 2 | 3 | E | 7 | 24980681 |
| | Congenital myopathy with central nuclei | 2 | Axial weakness respiratory involvment rare cardiomyopathy | 0 | Nocturnal ventilation / Cardiac monitoring | 2 | 3 | C | 7 | 24980681 |
| | Congenital myopathy with arthrogryposis multiple | 2 | Birth - pre natal<br>severe functional retraction involment | 0 | Symptomatic treatment, cardiac monitoring, orthopedic surgery | 0 | 1 | C | 3 | 24980681 |
| | TMD ("Tibial muscular dystrophy") | 2 | Early adult-onset, proximal involment with evolution | 0 | Eymptomatic treatment, cardiac monitoring | 1 | 3 | B | 6 | 24980681 |
| | Emery-Dreifuss muscular dystrophy-like | 2 | Early-onset cardiac involment and severe retraction | 0 | Nocturnal ventilation / Cardiac monitoring | 2 | 3 | A | 7 | 24980681<br>27854229<br>17444505 |
| | LGMDR10 | 3 | Severe proximal weaknes<br>respiratory involment | 0 | Cardiac monitoring / pacemaker | 2 | 3 | A | 8 | 15728284<br>20571043 |
| | TMD ("Tibial muscular dystrophy") | 1 | late adult-onset, tibialis anterior weakness. | 0 | symptomatic treatment, cardiac monitoring. | 1 | 3 | B | 2 | 29435569 |
| | HMERF | 3 | Adult onset (average age of approximately 35 years), distal or proximal muscle weakness, respiratory insufficiency around 50 years, rare cardiac involment. | 0 | Cardiac monitoring and non-invasive ventilation | 2 | 3 | B | 8 | 29435569<br>22577215<br>15728284<br>20571043 |
| | LGMD with cardiomyopathy (LGMDR10) | 2 | Variable proximal weakness and cardiomyotpathy | 0 | Nocturnal ventilation / Cardiac monitoring | 2 | 3 | A | 7 | 29435569<br>15728284<br>20571043<br>29691892<br>29139381 |
| VCP | Inclusion body myopathy with early-onset Paget disease and frontotemporal dementia 1<br>Charcot-Marie-Tooth disease, type 2Y | 2 | Adult onset, proximodistal muscle weakness, Paget's disease of the bone, dementia, respiratory failure | 0 | Paliative treatment | 1 | 3 | C | 6 | 18845250 |
| YARS2 | Myopathy, lactic acidosis, and sideroblastic anemia 2 | 3 | Childhood, progressive exercise intolerance, sideroblastic anaemia, lactic acidosis, mitochondrial muopathy, cardiomyopathy plus respiratory failure : early death | 0 | Symptomatic treatment | 0 | 0 | E | 3 | 20598274<br>24430573<br>30026338 |
| ZBTB42 | Lethal congenital contracture syndrome 6 | 3 | Fetal lethal form, IUCN with joint contractions, stomach absence, hydramnios | 0 | Symptomatic and palliative treatment | 0 | 0 | E | 3 | 25055871 |
| ZC4H2 | Wieacker-Wolff syndrome | 2 | Neonatal, arthrogrypose, muscle weakness --> +/- neonatal respiratory distress, dysmorphia, intellectual impairment + psychomotor retardation, camptodactylia, hip dislocation, scoliosis, foot bots, convulsion, spasticity. | 0 | Symptomatic and palliative treatment (orthopedic surgery, physiotherapy, ergotherapy). | 1 | 1 | C | 4 | 23623388<br>26056227<br>28345801 |
